# Household Transmission of SARS-CoV-2: Insights from a Population-based Serological Survey

**DOI:** 10.1101/2020.11.04.20225573

**Authors:** Qifang Bi, Justin Lessler, Isabella Eckerle, Stephen A Lauer, Laurent Kaiser, Nicolas Vuilleumier, Derek AT Cummings, Antoine Flahault, Dusan Petrovic, Idris Guessous, Silvia Stringhini, Andrew S. Azman, for the SEROCoV-POP Study Group

**Affiliations:** Department of Epidemiology, Johns Hopkins Bloomberg School of Public Health; Geneva Center for Emerging Viral Diseases and Laboratory of Virology, Geneva University Hospitals, Geneva, Switzerland; Department of Microbiology and Molecular Medicine, Faculty of Medicine, University of Geneva, Geneva, Switzerland; Division of Infectious Diseases, Geneva University Hospitals, Geneva, Switzerland; Department of Medicine, Faculty of Medicine, University of Geneva, Geneva, Switzerland; Division of Laboratory Medicine, Geneva University Hospitals, Geneva, Switzerland; Department of Biology, University of Florida, Gainesville, USA; Emerging Pathogens Institute, University of Florida, Gainesville, USA; Division of Tropical and Humanitarian Medicine, Geneva University Hospitals, Geneva, Switzerland; Department of Health and Community Medicine, Faculty of Medicine, University of Geneva, Geneva, Switzerland; Institute of Global Health, Faculty of Medicine, University of Geneva, Geneva, Switzerland; Division of Primary Care Medicine, Geneva University Hospitals, Geneva, Switzerland; University Centre for General Medicine and Public Health, University of Lausanne, Lausanne, Switzerland; Centre for Environment and Health, School of Public Health, Department of Epidemiology and Biostatistics, Imperial College London, London, UK

## Abstract

**Background:** Knowing the transmissibility of asymptomatic infections and risk of infection from household- and community-exposures is critical to SARS-CoV-2 control. Limited previous evidence is based primarily on virologic testing, which disproportionately misses mild and asymptomatic infections. Serologic measures are more likely to capture all previously infected individuals.

**Objective:** Estimate the risk of SARS-CoV-2 infection from household and community exposures, and identify key risk factors for transmission and infection.

**Design:** Cross-sectional household serosurvey and transmission model.

**Setting:** Geneva, Switzerland

**Participants:** 4,524 household members ≥5 years from 2,267 households enrolled April-June 2020.

**Measurements:** Past SARS-CoV-2 infection confirmed through IgG ELISA. Chain-binomial models based on the number of infections within households used to estimate the cumulative extra-household infection risk and infection risk from exposure to an infected household member by demographics and infector’s symptoms.

**Results:** The chance of being infected by a SARS-CoV-2 infected household member was 17.3% (95%CrI,13.7-21.7%) compared to a cumulative extra-household infection risk of 5.1% (95%CrI,4.5-5.8%). Infection risk from an infected household member increased with age, with 5-9 year olds having 0.4 times (95%CrI, 0.07-1.4) the odds of infection, and ≥65 years olds having 2.7 (95%CrI,0.88-7.4) times the odds of infection of 20-49 year olds. Working-age adults had the highest extra-household infection risk. Seropositive asymptomatic household members had 69.6% lower odds (95%CrI,33.7-88.1%) of infecting another household member compared to those reporting symptoms, accounting for 14.7% (95%CrI,6.3-23.2%) of all household infections.

**Limitations:** Self-reported symptoms, small number of seropositive kids and imperfect serologic tests.

**Conclusion:** The risk of infection from exposure to a single infected household member was more than three-times that of extra-household exposures over the first pandemic wave. Young children had a lower risk of infection from household members. Asymptomatic infections are far less likely to transmit than symptomatic ones but do cause infections.

**Funding Source:** Swiss Federal Office of Public Health, Swiss School of Public Health (Corona Immunitas research program), Fondation de Bienfaisance du Groupe Pictet, Fondation Ancrage, Fondation Privée des Hôpitaux Universitaires de Genève, and Center for Emerging Viral Diseases.

## Background

Household-centered studies provide an enumerable set of individuals known to be exposed to an infectious person, hence, they have played an important role for estimating key transmission properties of SARS-CoV-2. However, most published studies of SARS-CoV-2 household transmission rely on clinical disease (COVID-19), and/or PCR-based viral detection to identify infected individuals.^1,2^ Due to the narrow time window after exposure in which RT-PCR can be highly sensitive,^3^ case ascertainment based on virologic testing may miss infections, especially those that are mild or asymptomatic.^4^ This can lead to important biases and limit what can be studied, including underestimates of the importance of sub-clinical infections and household secondary attack rates.^4^

Serologic studies provide an alternative tool for understanding SARS-CoV-2 transmission. Serological tests remain sensitive to detecting past infections well beyond the period when the virus is detectable,^5–7^ thereby providing a measure of whether individuals have ever been infected.

Virologic and serologic studies have provided important insights into SARS-CoV-2 transmission. These include estimates of the household secondary attack rate (e.g., 17% in a recent meta-analysis^2^) and evidence of reduced infection rates among young children.^2,8,9^ However, in general, these estimates do not distinguish between intra- and extra-household transmission nor do they provide an estimate of transmission risk from a single infected individual. A notable exception is a household study from Guangzhou, China^10^, but this PCR-based study suffered from the limitations of virologic testing noted above. Hence, a number of critical gaps in the evidence remain, including the relative role of transmission between household members, the frequency of viral introductions into households from the community, the infectiousness of asymptomatic individuals, and the effect of age on transmission.

To help fill these gaps, we apply household transmission models to data from a cross-sectional, household-based population serosurvey of 4,534 people from 2,267 households in Geneva, Switzerland. We provide a serology-based assessment of transmission between intra- and extra-household contacts, identify risk factors for infection and transmission and estimate the relative risk of asymptomatic transmission. By doing so, we provide important evidence for guiding the COVID-19 pandemic response.

## Methods

### Study design, participants, and procedures

The SEROCoV-POP study is a cross-sectional population-based survey of former participants of an annual survey of individuals 20-74 years old representative of the population of Geneva (Canton), Switzerland. The full survey protocol is available online and a detailed description of the design and seroprevalence results were previously published.^11,12^

The SEROCoV-POP study invited all 10,587 participants of the previous annual surveys to participate in the study through email or post. Participants were invited to bring all members of their household aged 5 years and older to join the study. After providing informed written consent, participants either filled out a questionnaire online, in the days before their visit, or on site at the time of their visit. The questionnaire included questions about participants’ demographics, household composition, symptoms since January 2020, details on the frequency of extra-household contacts and reduction in social interaction since the start of pandemic. Only participants 14 years and older were asked about their frequency of extra-household contacts and changes in behavior. Despite this age cut off, we use more standard age cutoffs (10-19 years) in our analysis for comparability with other studies.^12^ We defined symptom presentation *a priori* as having reported any of: cough, fever, shortness of breath, or loss of smell or taste since January 2020 (symptoms reported in the 2-weeks prior to testing were excluded in a sensitivity analysis). We collected peripheral venous blood from each consenting participant. Households where all members provided blood samples were included in the present analysis (there was a 100% questionnaire response rate in this group). As blood was not collected from children under 5, all households with children in this age group were excluded. We conducted a sensitivity analysis with all households, regardless of whether all members provided blood samples, effectively treating household members outside the study as a community source of infection. The study was approved by the Cantonal Research Ethics Commission of Geneva, Switzerland (CER16-363).

### Laboratory analysis

We assessed anti-SARS-CoV-2 IgG antibodies in each participant using an ELISA (Euroimmun; Lübeck, Germany #EI 2606-9601 G) targeting the S1 domain of the spike protein of SARS-CoV-2; sera diluted 1:101 were processed on a EuroLabWorkstation ELISA (Euroimmun). An in-house validation study found that the manufacturer’s recommended cutoff for positivity (≥1.1) had a specificity of 99% and sensitivity of 93%, based on positive controls tested between 0 and 39 days after symptom onset. ^13^ In our primary analyses we defined seropositivity based on the cutoff recommended by the manufacturer and explored a higher cut-off of 1.5 (>1.5) in sensitivity analyses.^13^ As the presence of antibodies has been shown to be a reliable marker of past infection, we use the term ‘infected’ to refer to a seropositive individual.

### Statistical analyses

We fit chain binomial transmission models to estimate two primary quantities; the probability of extra-household infection from the start of the epidemic through the time of blood draw (referred to also as ‘community infections’ over the first epidemic wave) and the probability of being infected from a single infected household member over the course of his/her infectious period (referred to as ‘household exposures’).^14^ When fitting these models we explicitly consider all possible sequences of viral introductions to each household and subsequent transmission events within the household. For example, in a household with 2 seropositive individuals, both could have been infected outside of the household, or one could have been infected outside and then infected one other person within the household. We adapted models to estimate the within household and extra-household transmission risk according to the characteristics of potential infectees (age, sex, self-reported extra-household contact behavior) and, for within-household risk, those of the potential infectors (symptoms, age). We imputed missing data related to extra-household contacts based on household averages (see supplement). We simulate the proportion of infections attributable to extra-household and within household exposures.

We built a series of ten models including different combinations of individual-level characteristics (e.g., age, sex, self-reported contacts, symptoms) and compared their fit using the widely applicable information criterion (WAIC).^15^ We implemented the models in the Stan probabilistic programming language and used the *rstan* package (version 2.21.0) to sample from the posterior distribution and analyse outputs.^16^ We used weakly informative priors on all parameters to be normally distributed on the logit scale with mean of 0 and standard error of 1.5. We ran four chains of 1,000 iterations each with 250 warm-up iterations and assessed convergence visually and using the Gelman-Rubin Convergence Statistic (R-hat).^17^ All estimates are means of the posterior samples with the 2.5th and 97.5th percentiles of this distribution reported as the 95% credible interval. Full model and inference details are provided in the supplement and code needed to reproduce analyses are available at https://github.com/HopkinsIDD/serocovpop-households.

## Results

Between April 3rd and June 30th, 8,344 individuals coming from 4,393 households were successfully enrolled in the SEROCoV-POP study (Figure 1, Figure S1).^12^ The median enrollment date was May 22nd, 86 days after the first case was detected in Geneva (February 26, 2020). In 2,267 of these households, all members of the household were eligible, available, and provided a blood sample (4,354 individuals). The majority of these households were either one (37.9%, n=860) or two (39.2%, n=889) person households (Figure S2). The median household size in our study (2.0, interquartile range [IQR]=1,2) was similar to the general population in Geneva canton (median=2.0, IQR=1,3).^18^

**Figure 1.**
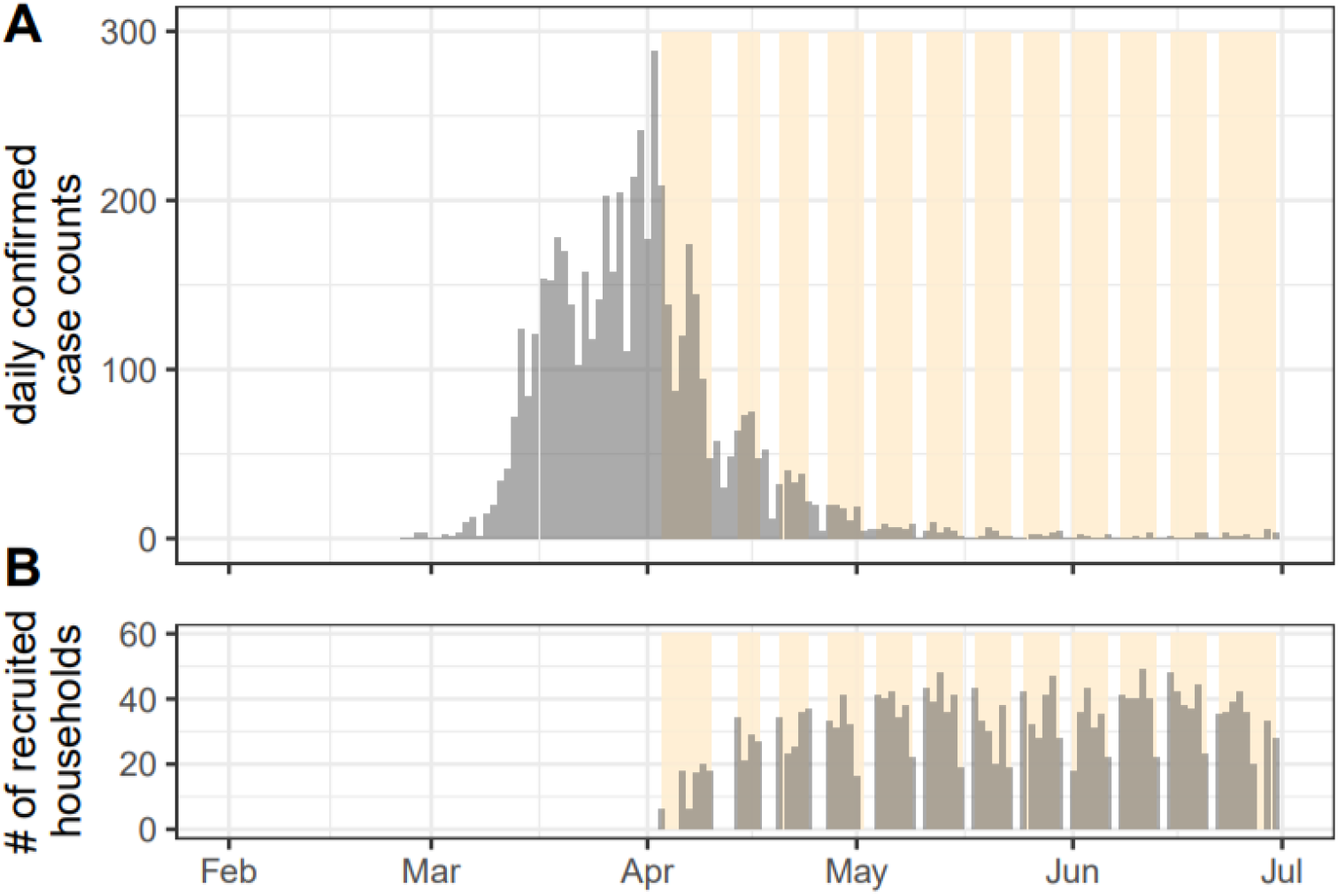
Epidemic curve and recruitment period of household serosurvey. (A) daily confirmed COVID-19 cases reported in Geneva up to July 1st, 2020. (B) Daily number of recruited households over the 12-week study period. First detected case in Geneva canton was reported on February 26th and the epidemic lasted about two months. Yellow bands indicate time periods of enrollment for each week. This includes all 4,438 households enrolled in the SEROCoV-POP study, not restricted to the complete households used in these analyses for which serostatus of all household members were available.

The median age of participants was 53 years (IQR=34,65), and 53.6% were female. Compared with the general canton population, our study sample included more individuals 50 years and older and fewer 20-49 year olds. Individuals in older age groups were more likely to live in smaller households: 94.6% (1,100/1,163) of people who were 65 years and older lived alone or in two-person households versus 44.5% (588/1,302) of those 20-49 years old (Table 1).

**Table 1.**
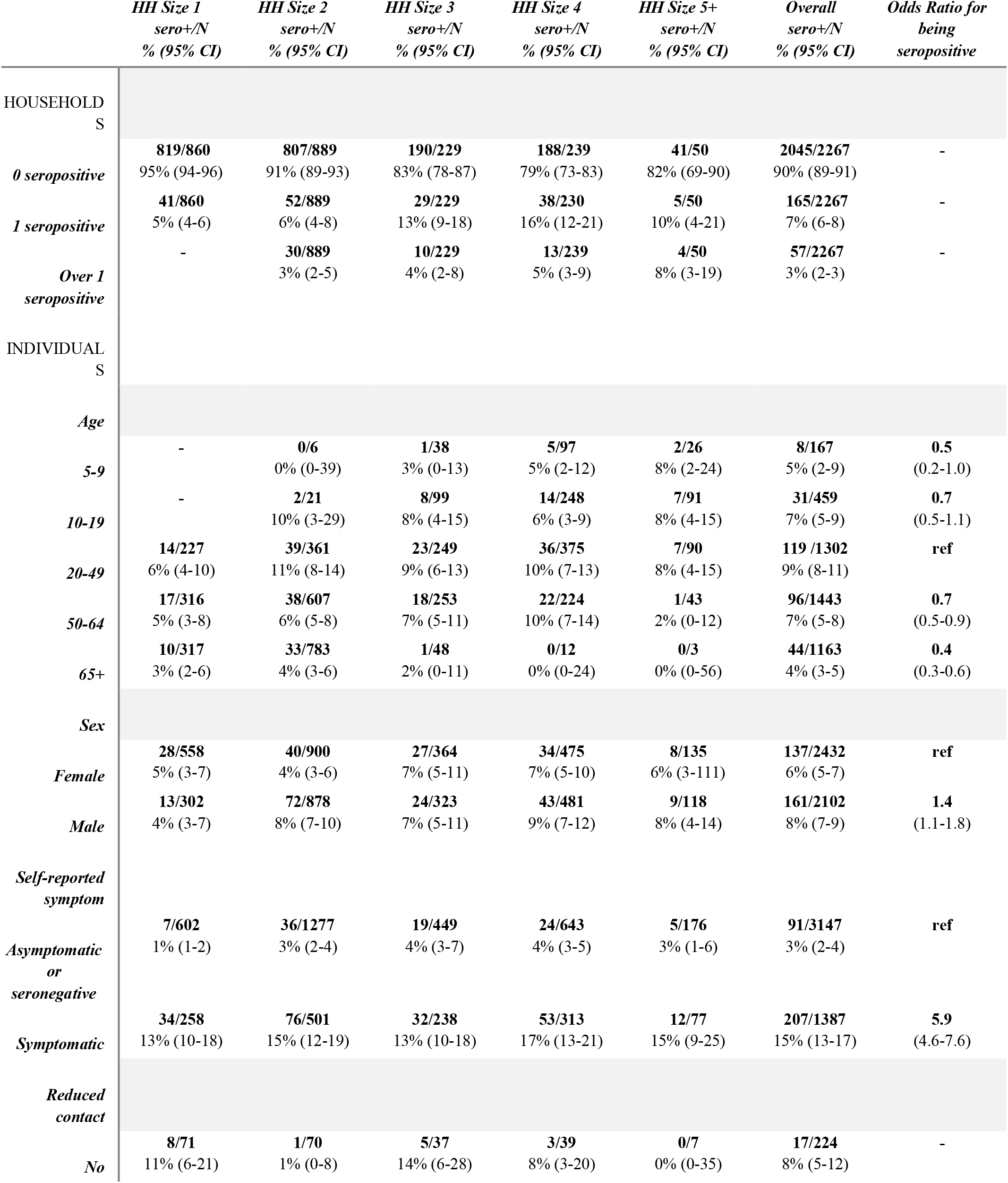
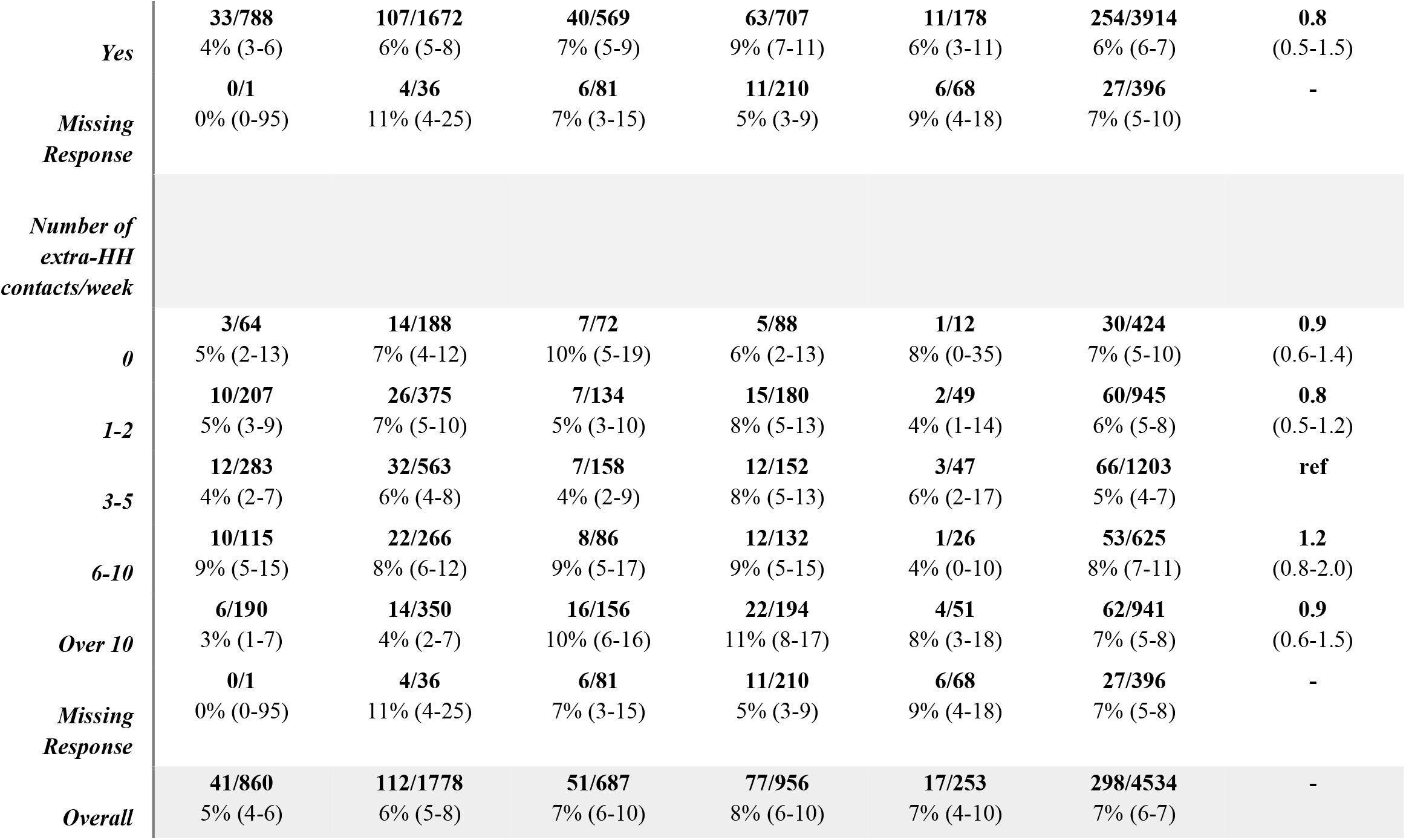
Number of recruited and seropositive individuals by age-group, sex and household size of the households they reside in.

Overall, 6.6% (298/4,534) of individuals tested positive for SARS-CoV-2 anti-S1 IgG antibodies by ELISA. Of the 2,267 households included in the analyses, 222 (9.8%) had at least one seropositive household member. The proportion of households with seropositive members increased from 4.8% (41/860) in households of size one, to 17.0% (39/229) in households of size three, and was relatively constant in larger households (Figure S2, Table 1, Figure S3). Symptoms consistent with COVID-19 were reported by 69.5% (207/298) of seropositive individuals although this was significantly lower in young children (37.5%, 3/8), similar to the results of an early modeling study.^19^

From the start of the epidemic in Geneva through the time of the serosurvey, the cumulative risk of infection from extra-household exposures was 5.1% (95% Credible Interval [CrI] 4.5-5.8%). The probability of being infected from a single infected household member was 17.3% (95% CrI 13.7-21.7%, Figure 2).

**Figure 2:**
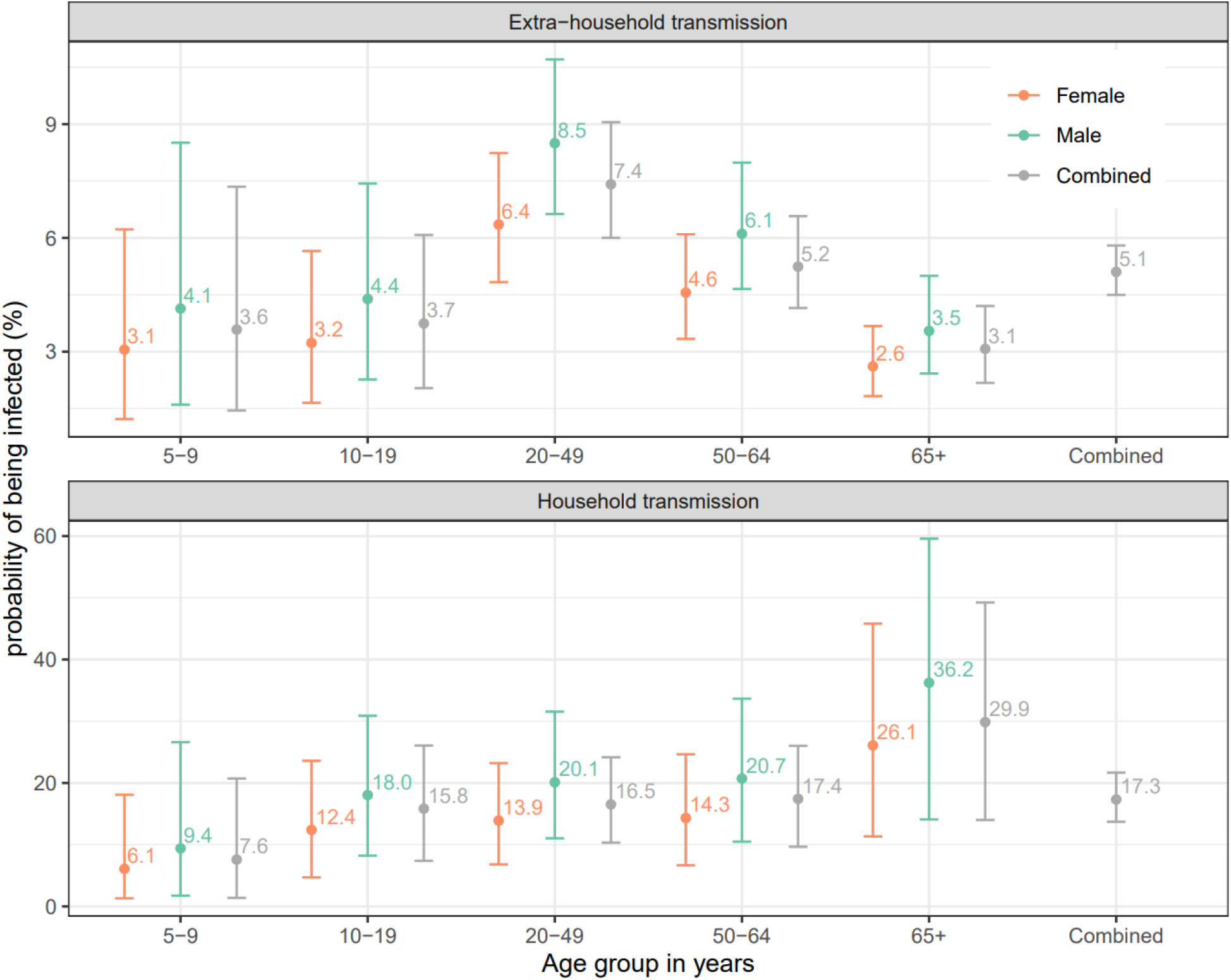
Estimated median probability of (A) extra-household infection from the start of the epidemic in Geneva until the time of the serosurvey and (B) infection from a single infected household member by age group and sex of the susceptibles. Bars represent 95% credible intervals. Probabilities of being infected by sex and age group of the exposed individuals are estimated by a model only including age and sex of the exposed individuals (model 2, orange/green bars; see Table S2). Probabilities of being infected by the age group of the exposed individuals combining males and females (left four grey bars on both panels) are estimated with an age-only model (model 1). The overall probabilities of being infected (rightmost grey bar on both panels) are estimated with the null model (model 0).

The risk of being infected by a household member increased with age (Figures 2,3). Compared to 20-49 years olds, 5-9 years olds had less than half the odds of being infected by an infected household member (OR=0.4, 95%CrI 0.1-1.4), while those 65 years and older had nearly three times the odds (OR=2.7, 95%CrI 0.9-7.4). Though credible intervals on individual estimates are wide, inclusion of age substantially improved model fit (ΔWAIC −14.8, Table S2). In contrast, the extra-household infection risk was the highest among working age adults (20-49 years olds). Compared to this group, 5-9 year olds (OR=0.3, 95%CrI 0.1-0.8) and those 65 years and older (OR=0.2, 95%CrI 0.1-0.9) had the lowest risk (Figure 2, Table S4). Models allowing for differential risk of transmission by the age of the infector were not well supported by the data (ΔWAIC −15.5 to −24.7) and included no significant differences between ages (Table S2).

**Figure 3.**
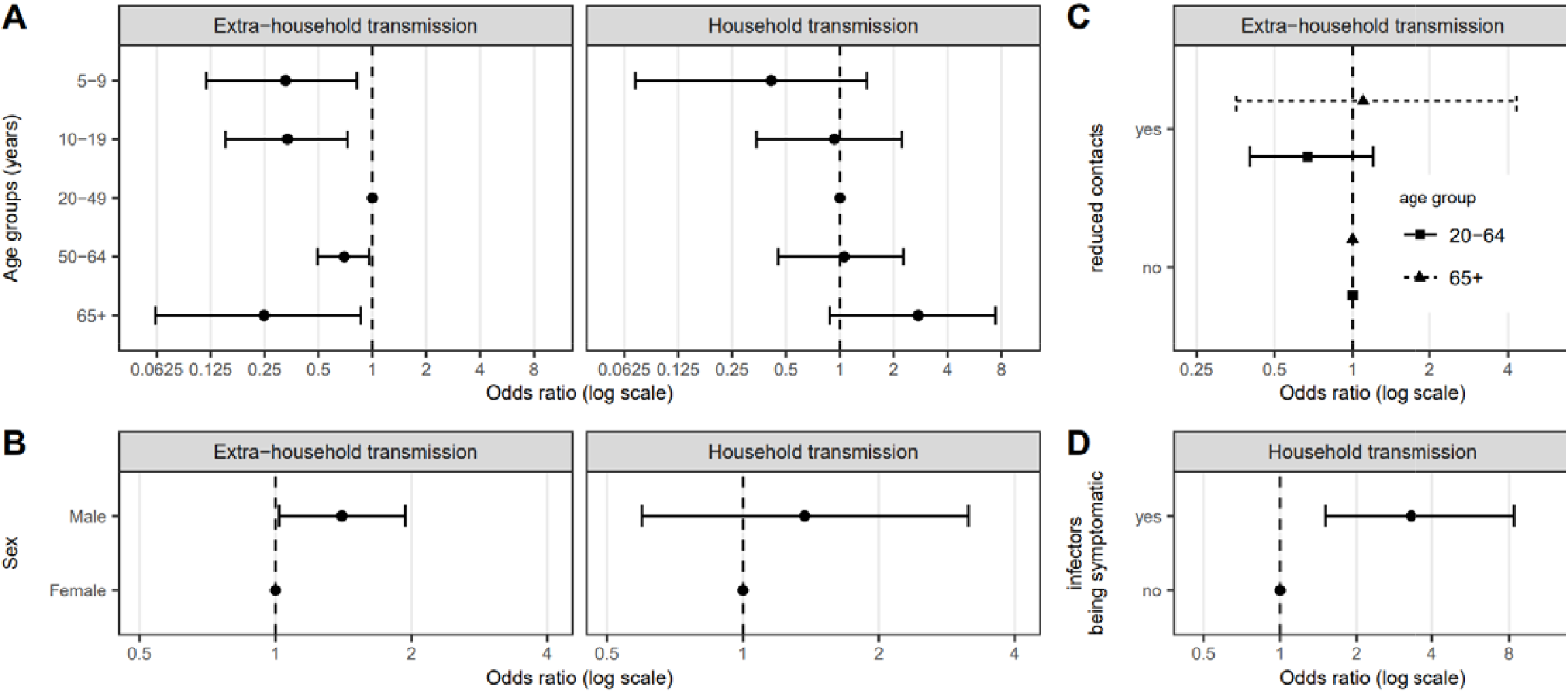
**Relative odds of being infected outside the household and** from a single infected household member by individual characteristics of the exposed individuals, A) age group, B) sex, C) self-reported reduction in social interaction since the start of the pandemic and D) potential infectors’ symptom status. Odds ratios and credible intervals, shown on the log-scale, are estimates from model 5 (See Table S2).

Males were more likely to be infected both outside (OR=1.4, 95%CrI 1.0-1.9) and inside (OR=1.4, 95%CrI 0.6-3.2) the household, though the latter estimate is less strongly supported by the data (Figure 3 & Table S2).

Seropositive household members not reporting symptoms had 0.30 times the odds (95%CrI: 0.12-0.67) of infecting another household member compared to those reporting symptoms consistent with COVID-19 (Figure 3). The difference was larger (OR=0.24, 95%CrI 0.10-0.56) when only considering those who reported symptoms more than two weeks before blood draw as symptomatic infections (Table S6).

We found some evidence that those aged 20-64 who reported reducing extra-household contacts during the pandemic had a reduced odds of extra-household infection (OR=0.67, 95%CrI 0.40-1.2). However, no similar reduction in the odds of infection was seen in those aged 65 or older and data were not available for younger individuals (Figure 3, Figure S5).

Using posterior distributions of parameters, we simulated the source of infection for all individuals in the study. We estimate that 22.5% (95%CrI 20.5-24.5) of all infections were caused by another household member, with the proportion of infections attributable to household transmission increasing with household size (Figure S8). In households with two individuals, 23.2% (95%CrI 19.6-26.8) of infections were between household members, increasing to 41.2% (95%CrI 29.4-47.1) in households of five people (Table S3). Of within-household infections, we estimate 14.7% (95%CrI 6.3-23.2) were due to individuals not reporting symptoms consistent with COVID-19.

Here we focus on the results of the best fitting models, but across the ten models considered (Table S2), estimates were qualitatively and quantitatively consistent with the primary findings. Similarly, we explored the sensitivity of our results to the ELISA seropositivity cutoff and found no qualitative differences in results (Figure S4).

## Discussion

The results presented here appropriately place symptomatic household transmission of SARS-CoV-2 in the context of community risk and asymptomatic spread. We show an approximate 1 in 6 risk (17.3%, 95%CrI 13.7-21.7%) of being infected by a single SARS-CoV-2 infected household member (Table S3). This contrasts with a 1 in 20 chance (5.1%, 95%CrI 4.5-5.8%) of being infected in the community over most of the first epidemic wave in Geneva, a period of roughly 2 months. Despite the high risk of transmission from an infected household member, as in many cities in high-income nations, households are mostly small limiting opportunities for onward transmission. Thus, less than a quarter (22.5%, 95%CrI 20.5-24.5%) of cases could be attributed to transmission between household members. While asymptomatic individuals appear to be less than a third (0.30, 95%CrI 0.12-0.66) as likely to transmit, they cannot be dismissed as inconsequential to disease spread, and are responsible for one in six (14.7%, 95%CrI 6.3-23.2) within-household transmissions in this study. Our results further illustrate the dual roles of biology and social behavior in shaping age-specific infection patterns, with the age signature of risk within households indicative of low biological susceptibility in the very young, and elevated susceptibility in the old; while extra-household risk seems more driven by behavior, with working age adults being at the highest risk.

It has long been thought that asymptomatic individuals are less likely to transmit than symptomatic ones, though studies have recovered similar concentrations of viral RNA from naso-pharyngeal samples from these two groups.^20^ By using serological data, we were able to show that those not reporting symptoms have one-third the odds of transmitting within households as symptomatic ones; and ultimately caused about 10% of household infections. This reduced transmissibility may be due to reduced duration of viral shedding and reduced ability to mechanically spread virions (e.g., through coughs). We did not assess the role of asymptomatics in community spread, but it is plausible that they may play an even larger role there, as symptomatic individuals are more likely to stay home or take extra precautions to reduce exposures when sick.

As with previous studies of SARS-CoV-2 transmission among household members and other close contacts^2,21^, we find evidence supporting a reduced risk of infection from household exposures among young children, and elevated risk of infection among those 65 or older. However, it is important to note that we only find this reduced risk among the youngest children in our study (5-9 year olds), while 10-19 year olds have a similar risk profile to working age adults. This is consistent with the hypothesis that young children may be biologically less susceptible to SARS-CoV-2 infection, though heterogeneity in social contact and other behaviors within households cannot be ruled out.

Patterns of extra-household infection suggest social factors dominate this risk, as both young children and older adults are at reduced risk of infection compared to working age adults. As children have returned to schools in Geneva, the social factors driving this pattern have likely changed significantly and we may see children become a more significant source of extra-household infections ^22^, despite their apparently lower susceptibility. The risk that infected young children pose to their household members is unclear; the sample size was likely too low to detect small to moderate differences in risk. We did not find any significant relationship between age of infector and probability of transmission (nor did including these terms improve model fit), but children are less often symptomatic^23^ and we did find a strong relationship between symptoms and transmission.

Our study has a number of important limitations. Symptoms were self-reported and, given that the times of infection are unknown, they may not necessarily have been a result of the SARS-CoV-2 infection.

We cannot exclude recall bias in symptom reports and other self-reported exposures. Further, we looked at only a narrow range of symptoms to increase specificity, which left out more general potentially SARS-CoV-2-related symptoms (e.g., nausea, diarrhea). We detected only eight seropositive children under the age of 10, leading to large uncertainty in age-specific risk estimates for this group. While validation data of the Euroimmun ELISA from across the world have confirmed its high specificity and sensitivity for detecting recent infections,^13,24,25^ most data are from adults, and it is possible that performance in young children may be different. Although most of the participants in the study were recruited after the epidemic peak, it is possible that we did not fully capture all infections in each household due to insufficient time to mount a detectable response or due to waning of responses. However, when conducting stratified analyses including households recruited early and late, we found few qualitative differences in the primary results (Figure S5). While we included only households where all household members provided blood samples in the main analysis, sensitivity analyses of all enrolled individuals led to similar primary results (Table S6).

This study captures infections that occurred during the first wave of the pandemic in Geneva, a period of time when workplaces and schools were largely closed and peoples’ social contacts were greatly reduced. In future phases of this pandemic, we may expect differences in our estimates of the proportion of transmission that occurs between household members. While we found no evidence in previous analyses of these data for differences in seropositivity by neighborhood wealth or education,^11^ these and other indicators of wealth might be associated with transmission risk within Geneva or in other populations. Likewise, the general nature of the Geneva population and the control measures in place may limit the generalizability of our estimates of absolute risk of infection, attributable fraction, and extra-household risks. For example, the increasing importance of household transmission with increasing household size (Figure S8), suggests household transmission would be far more important in settings with larger households. However, we believe our estimates of relative risks by age and symptom status within households, which are likely more biologically driven, should be generalizable to most settings; as should our general observations about how social and biological factors influence different types of transmission.

Our study highlights how biological and social factors combine to shape the risk of SARS-CoV-2 infection. While we expect some differences across settings, we believe that the general trend in per-exposure infection risk by age and sex and increased infectiousness of symptomatic individuals are fundamental attributes of this pandemic. These differences have important implications for guiding patient care and public health policy. For example, increased susceptibility of the oldest individuals suggests that rapid and aggressive measures are needed to protect them as soon as there is any possibility that SARS-CoV-2 was introduced into their living environment. At the population level, quantifying the infectiousness of asymptomatics can help us understand the extent the pandemic is driven by asymptomatic infections. Continued serological and virologic monitoring of diverse populations with detailed analyses like those presented here are critical to the continued evidence-based response to this pandemic.

## Data Availability

Code to reproduce analyses will be posted on github (https://github.com/HopkinsIDD/serocovpop-households) and data will be made available upon request to the corresponding author.

## Author Contributions

Drs Azman and Stringhini had full access to all of the data in the study and took responsibility for the integrity of the data and the accuracy of the data analysis. Drs Azman and Lessler have contributed equally.

*Concept and design*: Bi, Lessler, Stringhini, Azman

*Acquisition, analysis, or interpretation of data*: Bi, Lessler, Eckerle, Lauer, Kaiser, Vuilleumier, Cummings, Flahult, Petrovic, Gessous, Stringhini, Azman

*Drafting of the manuscript*: Bi, Lessler, Stringhini, Azman

*Critical revision of the manuscript for important intellectual content*: Bi, Lessler, Eckerle, Lauer, Kaiser, Vuilleumier, Cummings, Flahult, Petrovic, Gessous, Stringhini, Azman

*Statistical analysis*: Bi, Lessler, Lauer, Azman

*Obtained funding*: Stringhini, Gessous

*Administrative, technical, or material support*: Stringhini, Gessous, Petrovic, Kaiser, Vuilleumier

*Supervision*: Azman, Lessler, Stringhini, Kaiser

## Role of the Funder/Sponsor

The funders had no role in the design and conduct of the study; collection, management, analysis, and interpretation of the data; preparation, review, or approval of the manuscript; and decision to submit the manuscript for publication.

## Supplemental material

**Figure S1.**
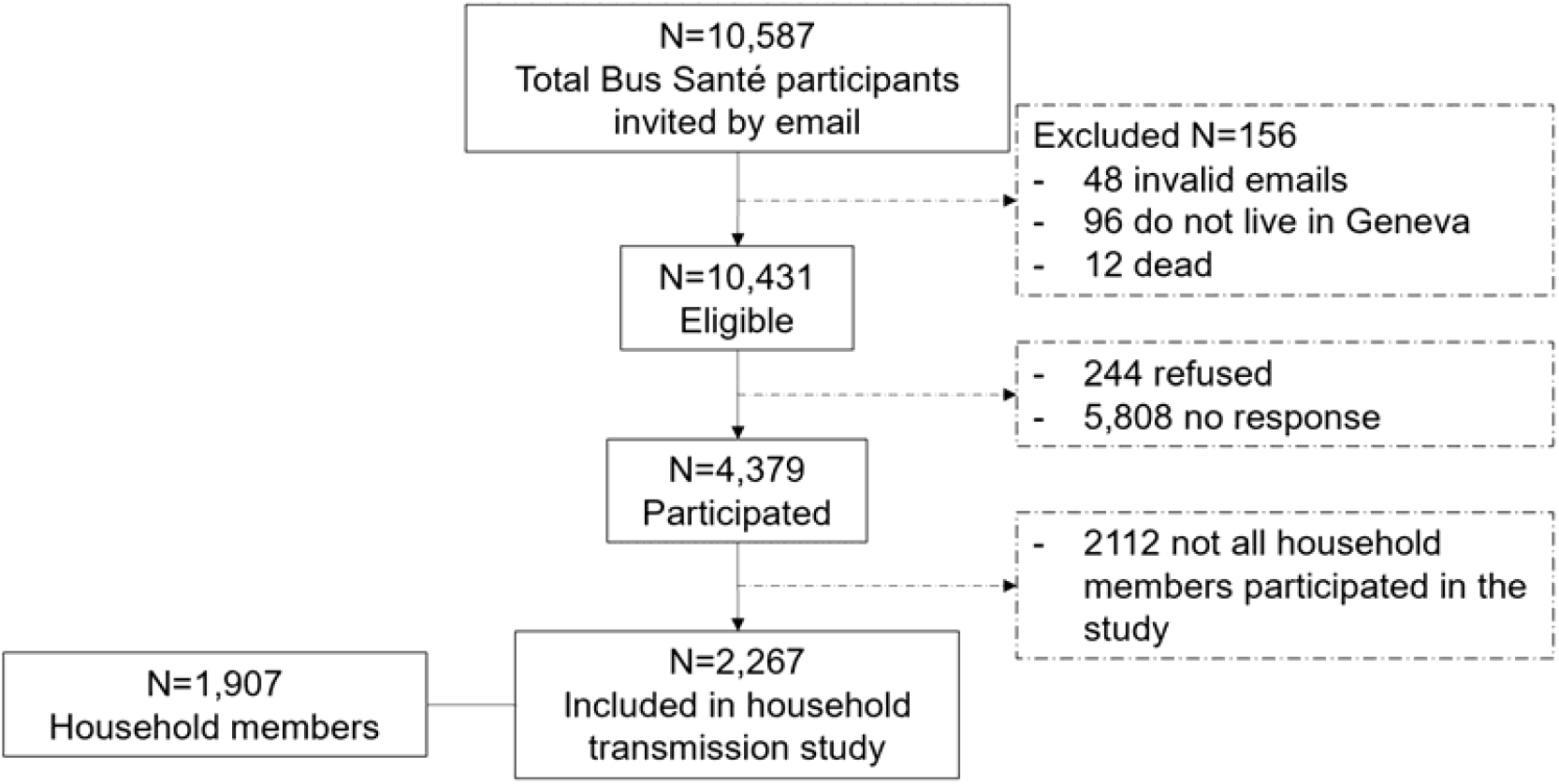
Study enrollment flowchart.

**Figure S2.**
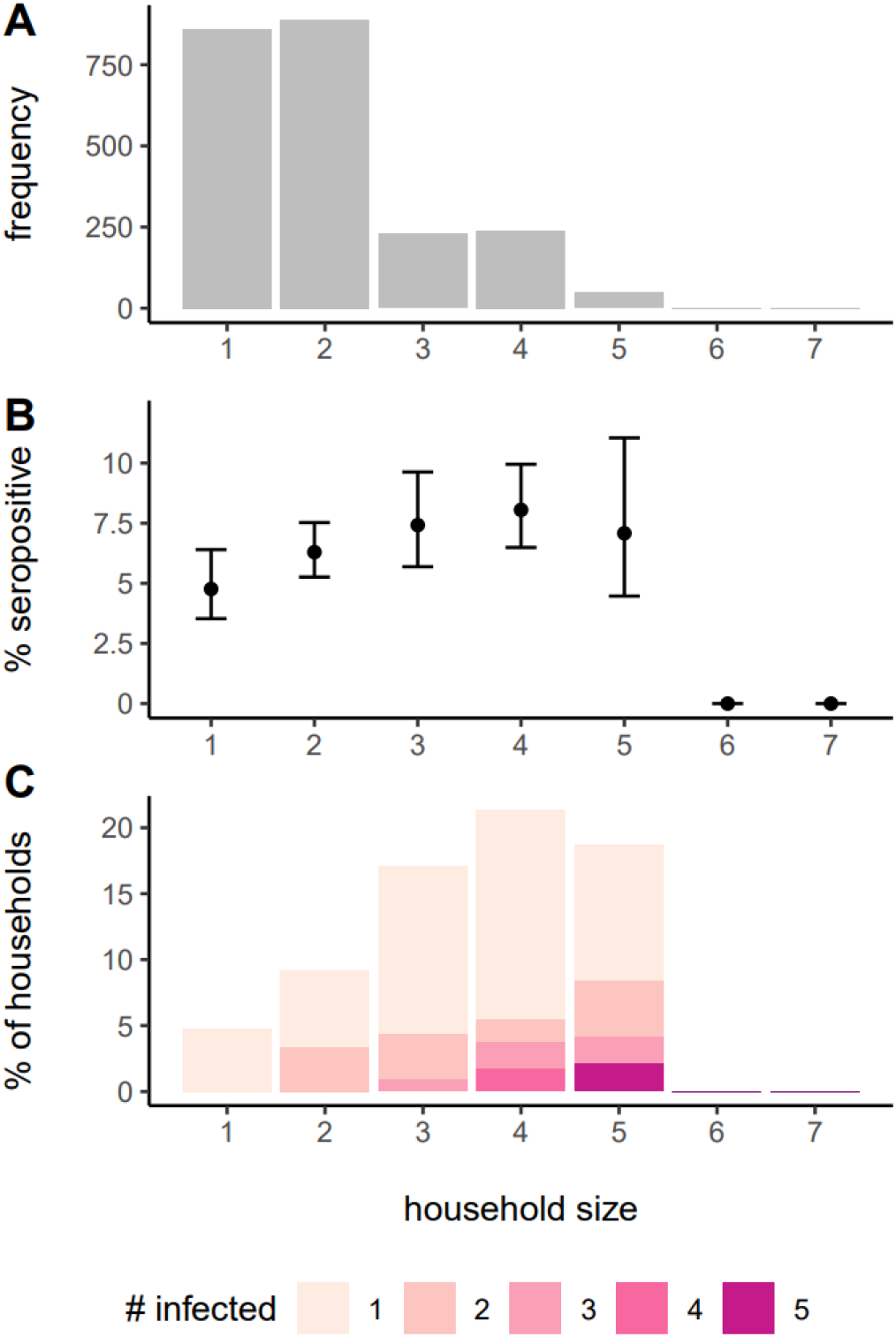
Frequency of households of different sizes in the study (A), proportion seropositive by household size (B) and distribution of the number of seropositive people in household by size (C).

**Figure S3.**
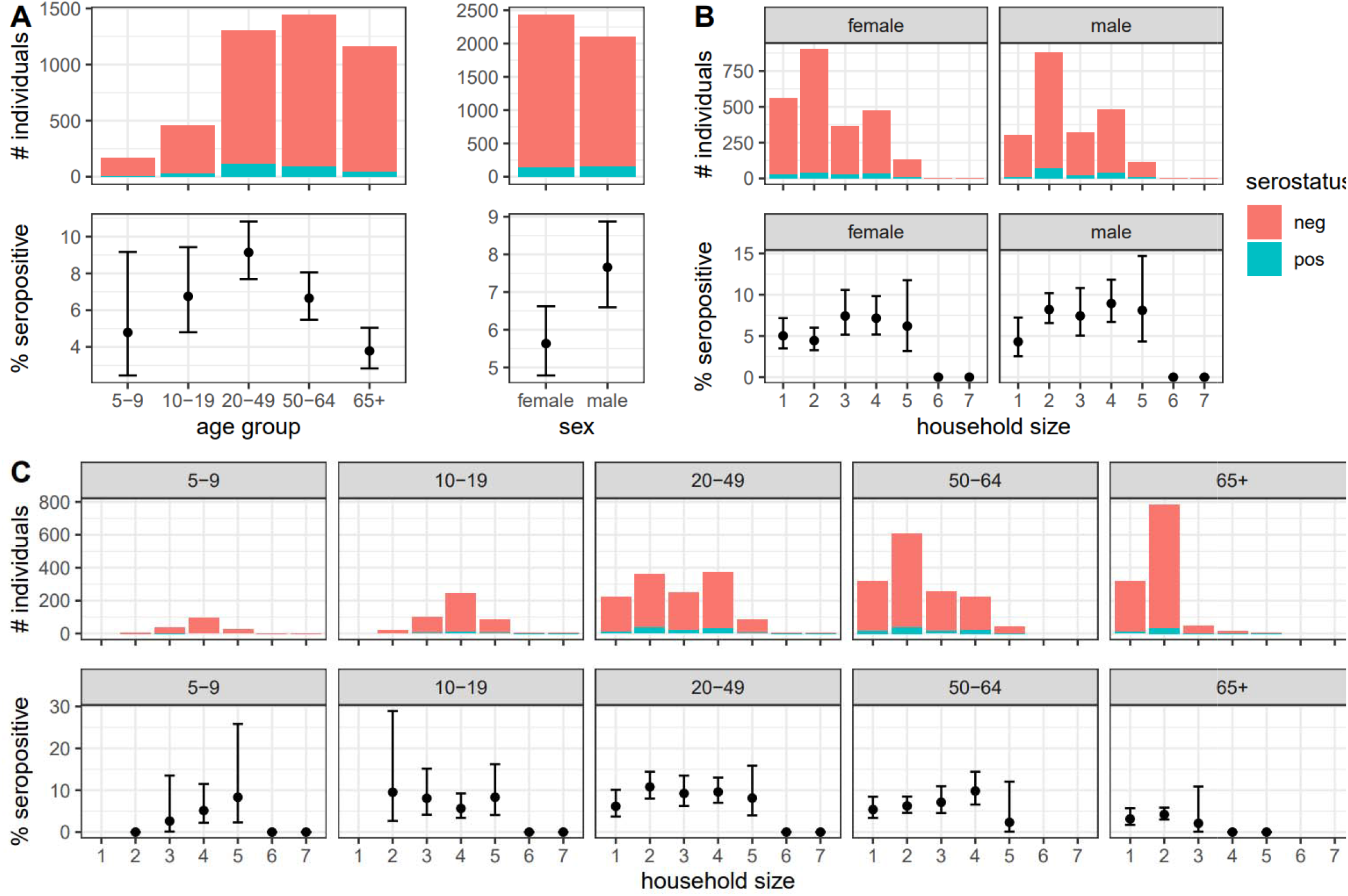
Study population by age group, sex, and household size. A) Number of individuals and seropositivity rate by age group, and by sex. B) Number of individuals and seropositivity rate by household size and sex. C) Number of individuals and seropositivity rate by household size and age group. Bar plots show the number of individuals in each group. Those tested seropositive and seronegative were colored in red and green respectively. The interval plots show seropositivity and accompanying 95% exact binomial confidence intervals in each group. Data that correspond to this figure are shown in Table 1.

**Figure S4.**
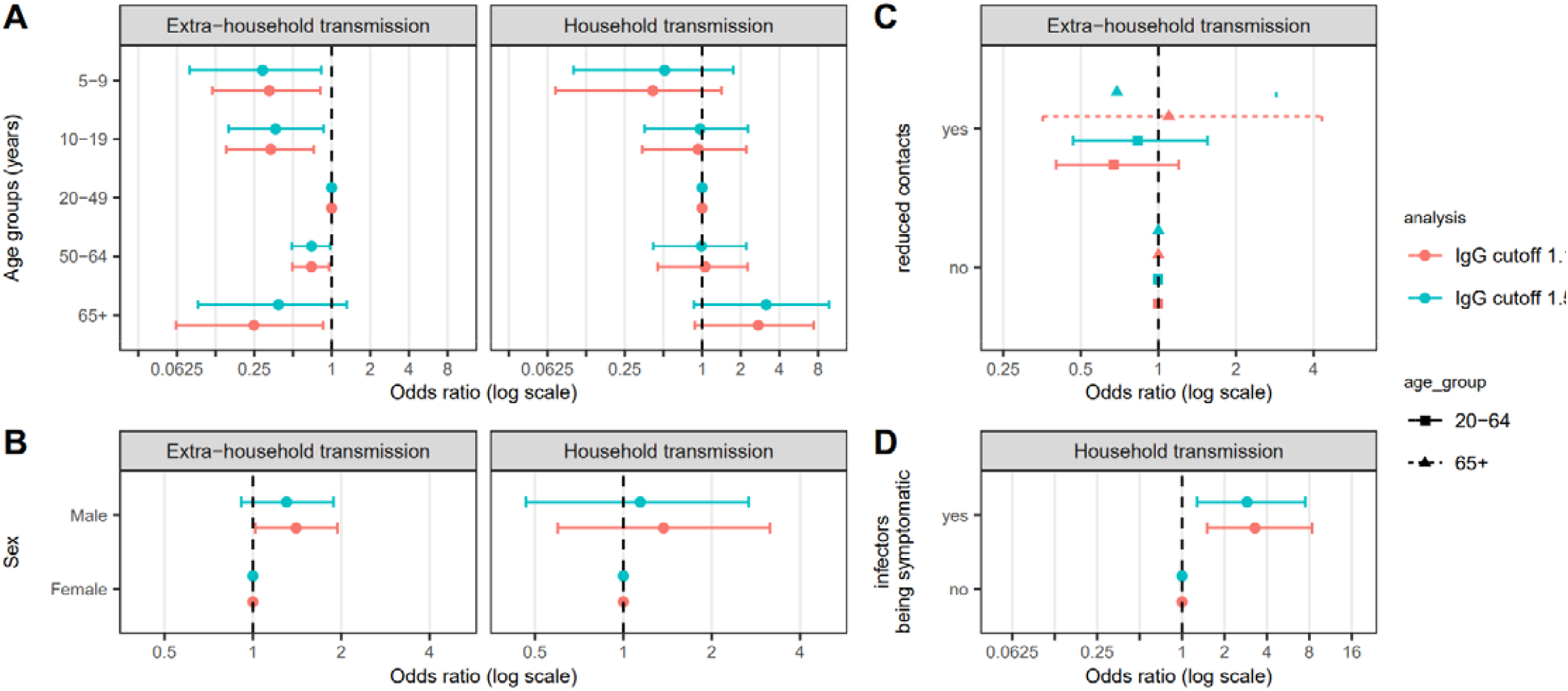
Relative odds of being infected (i.e., tested seropositive) by individual characteristics using different definitions of seropositivity. In the main analysis, all samples with an optical density to cutoff ratio ≥1.1 were classified as being seropositive. In the sensitivity analysis, all samples with an optical density to cutoff >1.5 were classified as being seropositive. An odds ratio greater than 1 indicates infection is more likely to occur in this group compared to the reference group. The reference group for the age-specific and sex-specific odds ratio are 20-49 years old and female, respectively. The reference group for symptom status of potential infectors was infectors being asymptomatic. The reference group for self-reported reduction in social interaction since the start of the outbreak was no reduction, and relative susceptibility was estimated by those 20-64 years old and those 65 years old and over separately.

**Figure S5.**
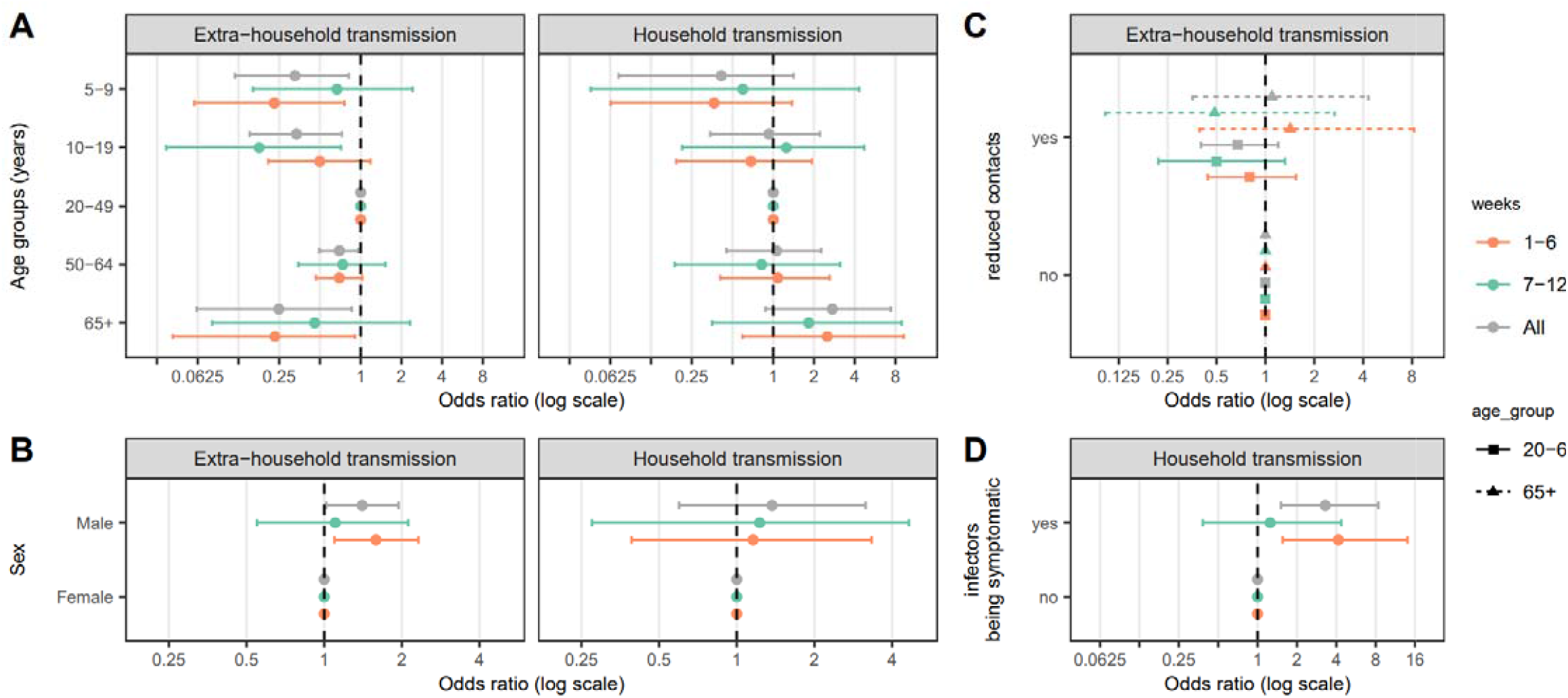
Relative odds of being infected by individual characteristics in the first and second half of the study period. First of the study period (first 6 weeks) spans from April 3rd to May 16th, and the second half (last 6 weeks) spans from May 18th to June 30th. An odds ratio greater than 1 indicates infection is more likely to occur in this group compared to the reference group. The reference group for the age-specific and sex-specific odds ratio are 20-49 years old and female, respectively. The reference group for symptom status of potential infectors was infectors being asymptomatic. The reference group for self-reported reduction in social interaction since the start of the outbreak was no reduction and was estimated for those 20-64 years old and those 65 years old and over separately.

**Figure S6.**
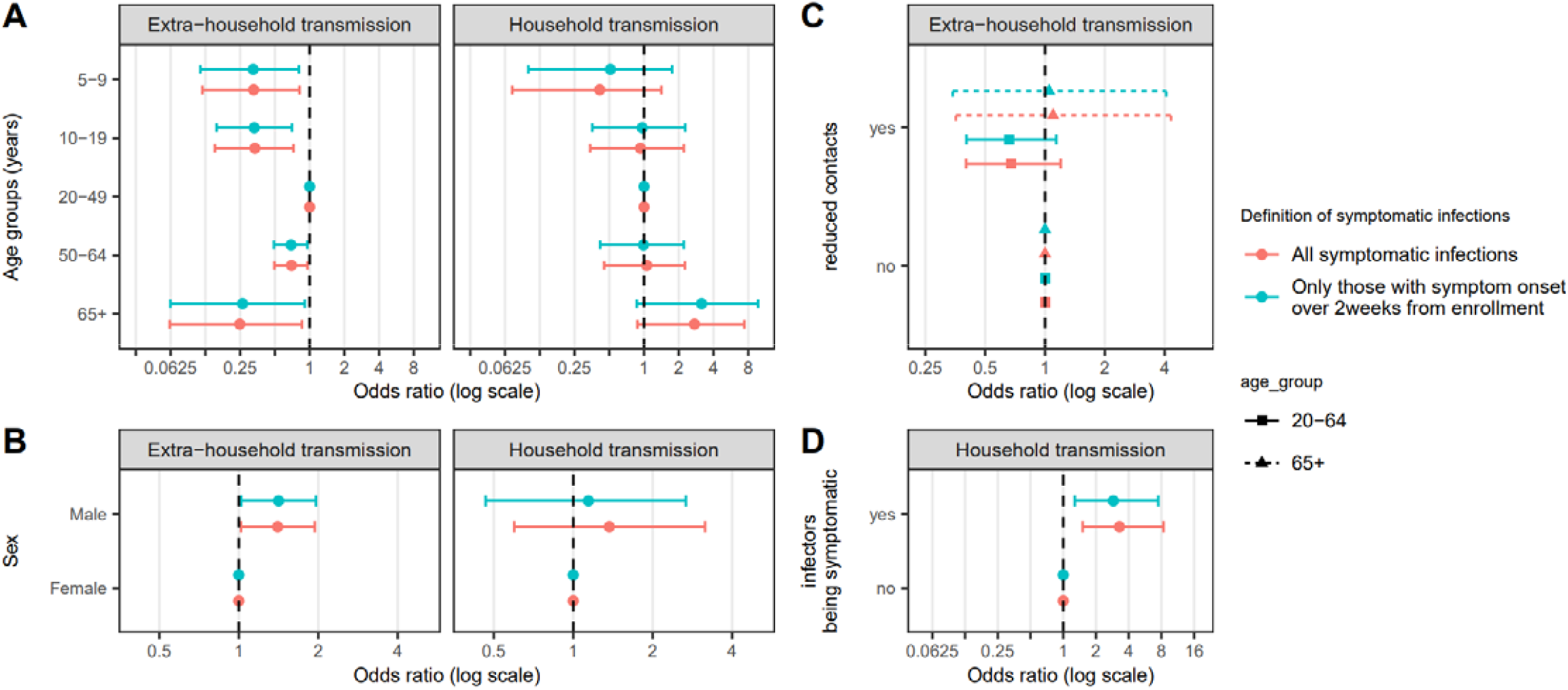
Relative odds of being infected by individual characteristics by different definitions of symptomatic cases. For sensitivity analyses, only seropositive individuals reporting symptoms more than two weeks before enrollment were considered symptomatic. An odds ratio greater than 1 indicates infection is more likely to occur in this group compared to the reference group. The reference group for the age-specific and sex-specific odds ratio are 20-49 years old and female, respectively. The reference group for symptom status of potential infectors was infectors being asymptomatic. The reference group for self-reported reduction in social interaction since the start of the outbreak was no reduction and was estimated for those 20-64 years old and those 65 years old and over separately.

**Figure S7.**
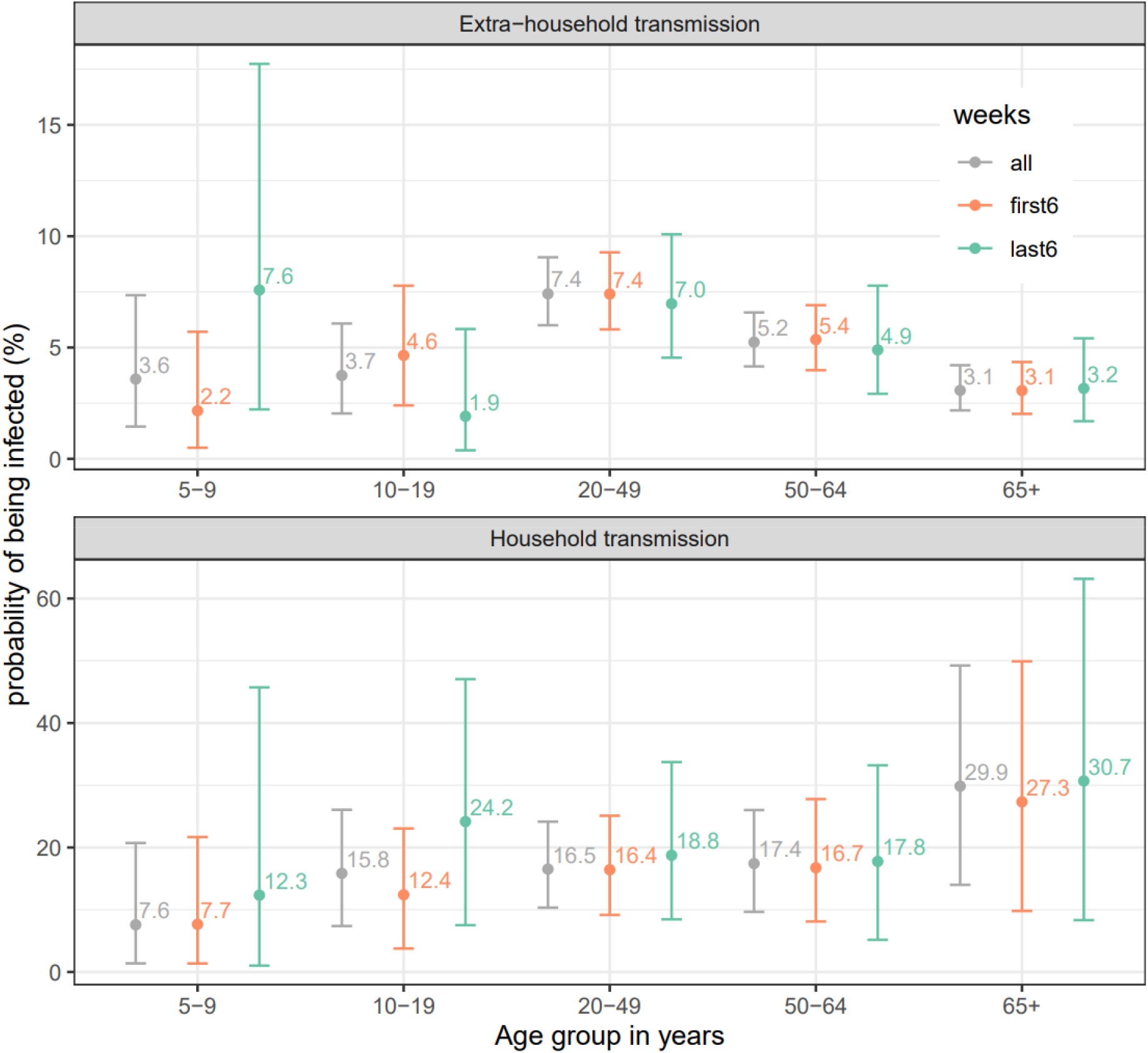
Median probability of (A) extra-household infection over the duration of the outbreak and (B) infection from a single infected household member by age group and sex of the susceptibles. Bars represent 95% credible intervals.

**Figure S8.**
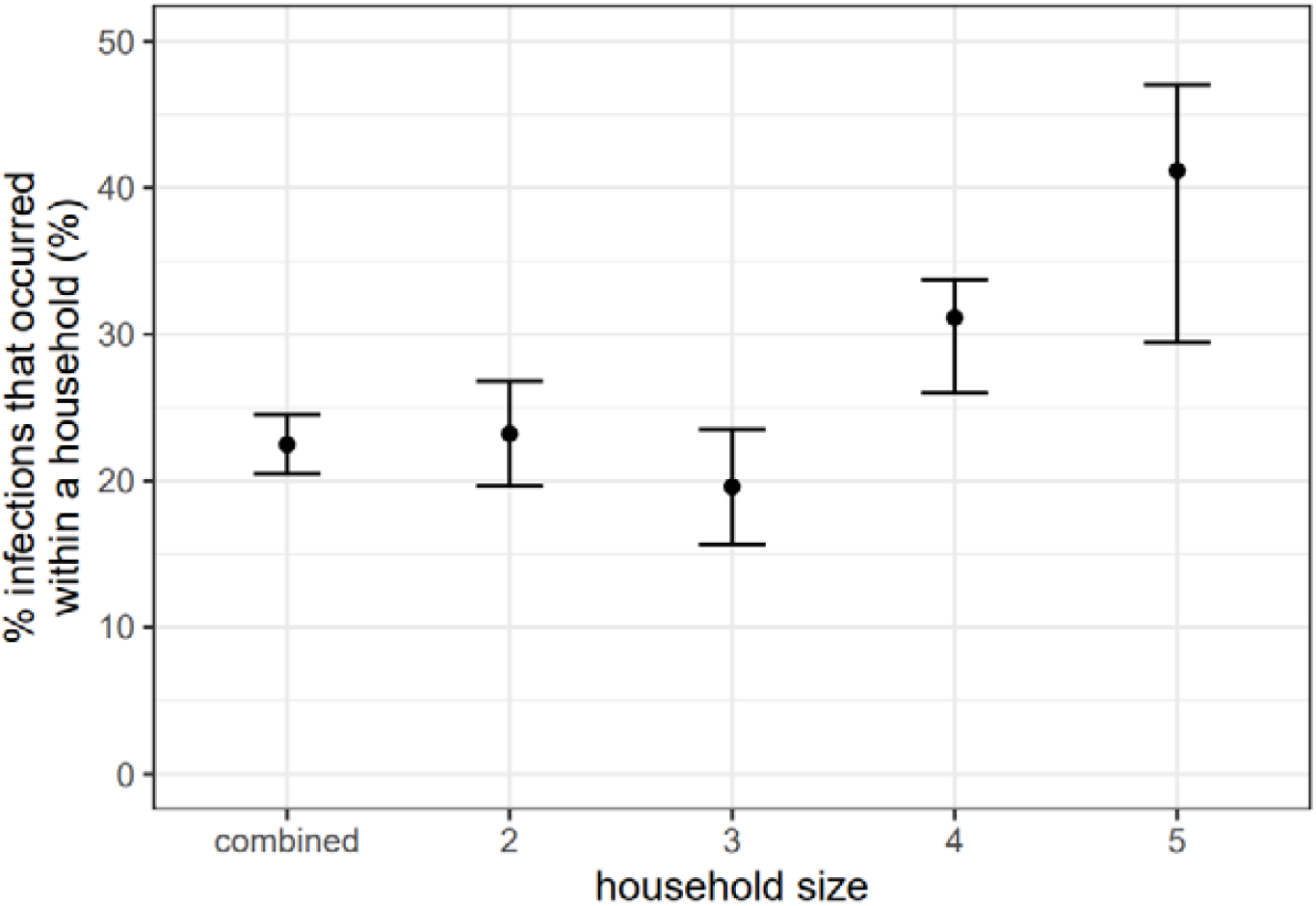
Proportion of infections that occurred within a household of various household size (median and 95% credible intervals).

**Figure S9.**
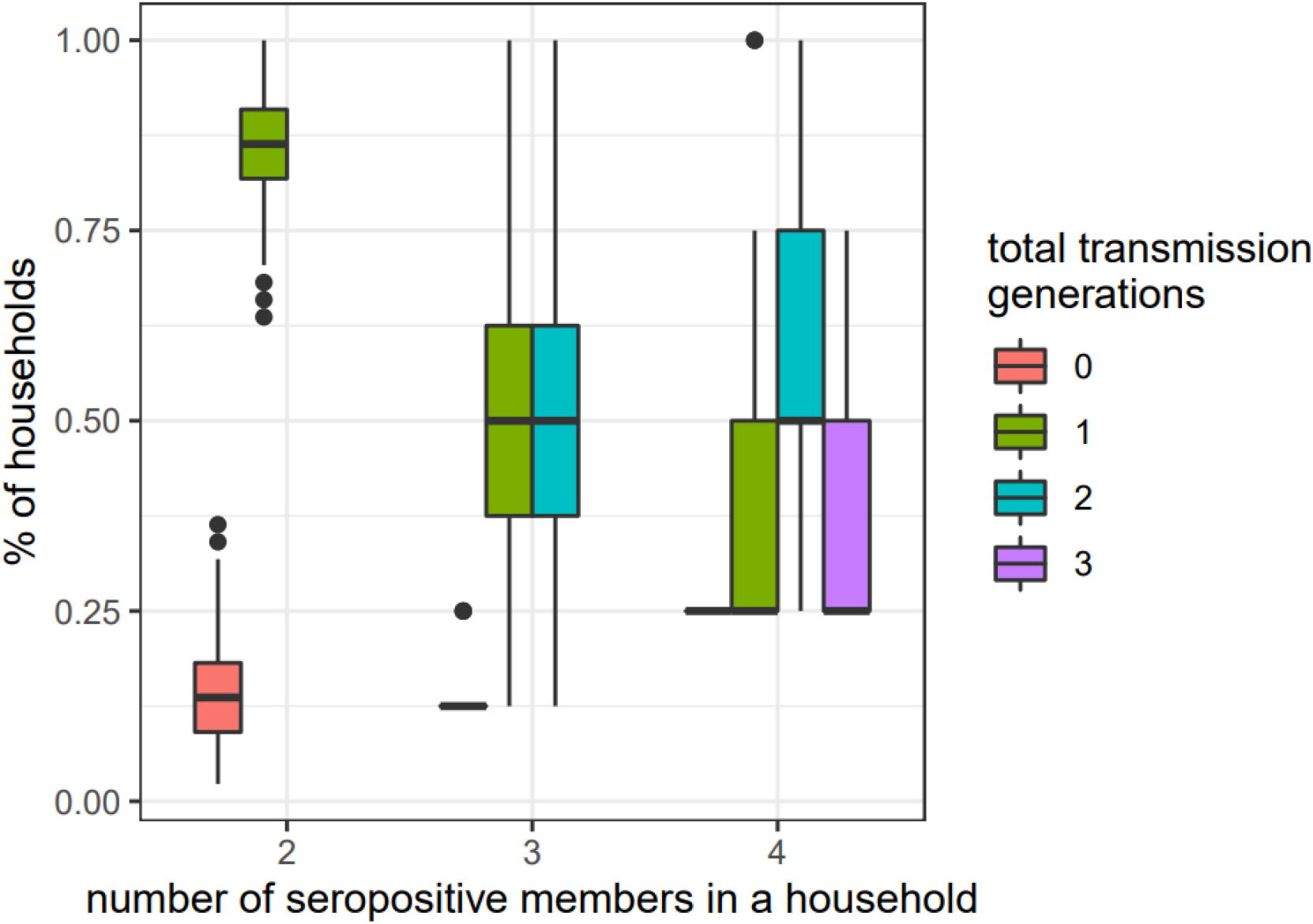
Distribution of the number of transmission generations within households by number of seropositives in a household. Zero generations means that all members were infected outside of a household. One generation indicates that there is one generation of transmission within a household cause from one or more index cases. There are 44 households with two seropositive households members (8 households with 3 seropositives and 4 households with 4 seropositives).

**Supplemental Table 1.** Household composition over the study period. HH stands for household. IQR stands for interquartile range.

| week | total households | % HHs with <10 years old (n) | % HHs with >65yrs old (n) | % HHs size = 1 (n) | average HH size (IQR) |
| --- | --- | --- | --- | --- | --- |
| 1 | 85 | 11.8% (10) | 18.8% (16) | 30.6% (26) | 2.33 (1, 3) |
| 2 | 111 | 5.4% (6) | 27.9% (31) | 29.7% (33) | 2.19 (1, 3) |
| 3 | 155 | 3.2% (5) | 32.9% (51) | 41.9% (65) | 1.95 (1, 2) |
| 4 | 153 | 9.8% (15) | 28.1% (43) | 33.3% (51) | 2.12 (1, 3) |
| 5 | 217 | 8.8% (19) | 25.8% (56) | 38.7% (84) | 2.07 (1, 3) |
| 6 | 226 | 4.9% (11) | 31.9% (72) | 35% (79) | 2.04 (1, 2) |
| 7 | 183 | 9.8% (18) | 31.1% (57) | 25.1% (46) | 2.33 (1.5, 3) |
| 8 | 218 | 2.3% (5) | 39.4% (86) | 45% (98) | 1.83 (1, 2) |
| 9 | 185 | 6.5% (12) | 40.5% (75) | 39.5% (73) | 1.98 (1, 2) |
| 10 | 232 | 4.7% (11) | 36.2% (84) | 37.9% (88) | 1.92 (1, 2) |
| 11 | 232 | 3.4% (8) | 47.4% (110) | 40.1% (93) | 1.81 (1, 2) |
| 12 | 270 | 4.4% (12) | 48.5% (131) | 45.9% (124) | 1.84 (1, 2) |
| Total | 2267 | 5.8% (132) | 35.8% (812) | 37.9% (860) | 2 (1, 2) |

**Supplemental Table 2.** Model performance and estimated parameters of the adapted chain binomial models that incorporate key individual-level factors (e.g., age, sex, reduced extra-household exposure, extra-household contact frequency of the exposed individuals and symptom status of the potential infectors) that may be associated with risk of infection from extra-household sources and by a single infected houseohld member. Odds ratio over 1 indicates higher risk of infection. CrI stands for credible interval.

|  | Model 1 |  | Model 2 |  | Model 3 |  |
| --- | --- | --- | --- | --- | --- | --- |
|  | age |  | age + sex |  | age/sex interaction |  |
|  | extra-household | household | extra-household | household | extra-household | household |

| Category ☐ | Odds Ratio<br>(95% CrI) | Odds Ratio<br>(95% CrI) | Odds Ratio<br>(95% CrI) | Odds Ratio<br>(95% CrI) | Odds Ratio<br>(95% CrI) | Odds Ratio<br>(95% CrI) |
| --- | --- | --- | --- | --- | --- | --- |
| **Age** |  |  |  |  |  |  |
| 5-9 | 0.5 (0.2, 1.0) | 0.4 (0.1, 1.4) | 0.5 (0.2, 1.0) | 0.4 (0.1, 1.5) | 0.4 (0.1, 1.1) | 0.3 (0.0, 1.4) |
| 10-19 | 0.5 (0.2, 0.9) | 0.9 (0.3, 2.3) | 0.5 (0.2, 0.9) | 0.9 (0.3, 2.1) | 0.4 (0.1, 0.9) | 1.4 (0.4, 4.6) |
| 20-49 | ref | ref | ref | ref | ref | ref |
| 50-64 | 0.7 (0.5, 1.0) | 1.1 (0.5, 2.3) | 0.7 (0.5, 1.0) | 1.0 (0.4, 2.4) | 0.6 (0.4, 1.0) | 0.8 (0.2, 2.6) |
| 65+ | 0.4 (0.3, 0.6) | 2.2 (0.7, 5.6) | 0.4 (0.3, 0.6) | 2.2 (0.8, 5.9) | 0.3 (0.2, 0.6) | 1.1 (0.2, 4.3) |
| **Sex** |  |  |  |  |  |  |
| Male | - | - | 1.4 (1.0, 1.9) | 1.6 (0.6, 3.9) | 1.1 (0.7, 1.7) | 1.6 (0.5, 5.2) |
| Female | - | - | ref | ref | ref | ref |
| **Age-sex**<br>**interaction** |  |  |  |  |  |  |
| 5-9 Male | - | - | - | - | 1.3 (0.3, 6.1) | 1.7 (0.2, 13.1) |
| 10-19 Male | - | - | - | - | 1.4 (0.5, 4.7) | 0.4 (0.1, 2.1) |
| 20-49 Male | - | - | - | - | ref | ref |
| 50-64 Male | - | - | - | - | 1.3 (0.7, 2.6) | 1.7 (0.2, 13.1) |
| 65+ Male | - | - | - | - | 1.5 (0.7, 3.5) | 5.2 (0.7, 43.7) |
| **ΔWAIC\*** | -14.8 |  | -20.2 |  | -19.2 |  |

\* WAIC of the null model (model 0) not model B or Q as a function of individual level characteristics is 2006.9.

Change in WAIC is calculated relative to WAIC of the null model. ☐ Individual characteristics refer to the characteristics of the susceptibles if not specified
|  | Model 4 |  | *Model 5\*\** |  | Model 6 |  |
| --- | --- | --- | --- | --- | --- | --- |
|  | **age + sex + infectors'**<br>**symptom** |  | **age + sex + infectors'**<br>**symptom + reduced contact** |  | **age + sex + infectors'**<br>**symptom + reduced contact**<br>**+ extra HH contacts** |  |
|  | **extra-**<br>**household** | **household** | **extra-**<br>**household** | **household** | **extra-**<br>**household** | **household** |
| Category ☐ | Odds Ratio<br>(95% CrI) | Odds Ratio<br>(95% CrI) | Odds Ratio<br>(95% CrI) | Odds Ratio<br>(95% CrI) | Odds Ratio<br>(95% CrI) | Odds Ratio<br>(95% CrI) |
| **Age** |  |  |  |  |  |  |
| 5-9 | 0.5 (0.2, 0.9) | 0.4 (0.1, 1.6) | 0.3 (0.1, 0.8) | 0.4 (0.1, 1.4) | 0.4 (0.1, 1.3) | 0.4 (0.1, 1.4) |
| 10-19 | 0.5 (0.2, 0.8) | 1.0 (0.4, 2.3) | 0.3 (0.2, 0.7) | 0.9 (0.3, 2.2) | 0.4 (0.1, 1.2) | 0.9 (0.3, 2.1) |
| 20-49 | ref | ref | ref | ref | ref | ref |
| 50-64 | 0.7 (0.5, 0.9) | 1.1 (0.5, 2.3) | 0.7 (0.5, 1.0) | 1.1 (0.5, 2.3) | 0.7 (0.5, 1.0) | 1.0 (0.4, 2.2) |
| 65+ | 0.4 (0.3, 0.6) | 2.7 (0.9, 7.9) | 0.2 (0.1, 0.9) | 2.7 (0.9, 7.4) | 0.3 (0.0, 1.5) | 2.8 (1.0, 7.5) |
| **Sex** |  |  |  |  |  |  |
| Male | 1.4 (1.0, 2.0) | 1.4 (0.6, 3.1) | 1.4 (1.0, 1.9) | 1.4 (0.6, 3.2) | 1.4 (1.0, 1.9) | 1.3 (0.6, 3.0) |
| Female | ref | ref | ref | ref | ref | ref |
| **Symptomatic (infector)** |  |  |  |  |  |  |
| yes | - | 3.3 (1.5, 8.9) | - | 3.3 (1.5, 8.4) | - | 3.4 (1.5, 8.5) |
| no |  | ref |  | ref |  | ref |
| **Reduced contact among 20-64 yo** |  |  |  |  |  |  |
| yes | - | - | 0.7 (0.4, 1.2) | - | 0.7 (0.4, 1.2) | - |
| no |  |  | ref |  | ref |  |
| **Reduced contact among 65+ yo** |  |  |  |  |  |  |
| yes | - | - | 1.1 (0.4, 4.3) | - | 1.1 (0.3, 5.7) | - |
| no |  |  | ref |  | ref |  |
| **Extra household contacts among 20-64 yo** |  |  |  |  |  |  |
| 0 | - | - | - | - | 1.6 (0.9, 2.9) | - |
| 1 to 2 | - | - | - | - | 1.2 (0.7, 2.0) | - |
| 6 to 10 | - | - | - | - | 1.8 (1.1, 3.0) | - |
| over 10 | - | - | - | - | 1.2 (0.7, 1.9) | - |
| **Extra household contacts among 65+ yo** |  |  |  |  |  |  |
| 0 | - | - | - | - | 0.5 (0.1, 1.8) | - |
| 1 to 2 | - | - | - | - | 1.6 (0.7, 4.1) | - |
| 6 to 10 | - | - | - | - | 2.0 (0.8, 5.2) | - |
| over 10 | - | - | - | - | 0.4 (0.1, 1.7) | - |
| **ΔWAIC** | -29.6 |  | -29.4 |  | -30.0 |  |

\*\* Main model of the study; OR estimates of model 5 are presented in Figure 4.

□ Individual characteristics refer to the characteristics of the susceptibles if not specified
|  | Model 7 |  | Model 8 |  | Model 9 |  |
| --- | --- | --- | --- | --- | --- | --- |
|  | age + sex + infectors' symptom + infectors' age |  | sex + symptom + infectors' age |  | age + sex + infectors' age |  |
|  | extra-household | household | extra-household | household | extra-household | household |
| Category □ | Odds Ratio (95% CrI) | Odds Ratio (95% CrI) | Odds Ratio (95% CrI) | Odds Ratio (95% CrI) | Odds Ratio (95% CrI) | Odds Ratio (95% CrI) |
| **Age of infectees** |  |  |  |  |  |  |
| 5-9 | 0.4 (0.2, 1.0) | 0.4 (0.1, 1.5) | - | - | 0.5 (0.2, 1.0) | 0.5 (0.1, 1.6) |
| 10-19 | 0.6 (0.2, 1.1) | 0.7 (0.2, 2.2) | - | - | 0.5 (0.2, 1.1) | 0.7 (0.1, 2.3) |
| 20-49 | ref | ref | - | - | ref | ref |
| 50-64 | 0.7 (0.5, 1.0) | 1.0 (0.4, 2.5) | - | - | 0.7 (0.5, 1.0) | 0.9 (0.3, 2.2) |
| 65+ | 0.4 (0.3, 0.6) | 1.8 (0.4, 9.0) | - | - | 0.4 (0.3, 0.6) | 2.1 (0.4, 8.0) |
| **Sex** |  |  |  |  |  |  |
| Male | 1.4 (1.0, 2.0) | 1.3 (0.5, 3.2) | 1.5 (1.1, 2.1) | 1.0 (0.5, 2.4) | 1.4 (1.0, 1.9) | 1.6 (0.7, 4.0) |
| Female | ref | ref | ref | ref | ref | ref |
| **Symptomatic (infector)** |  |  |  |  |  |  |
| yes | - | 3.6 (1.5, 10.4) | - | 3.9 (1.7, 10.6) | - | - |
| no | - | ref |  | ref | - | - |
| **Age of infectors** |  |  |  |  |  |  |
| 5-9 | - | 0.7 (0.0, 4.7) | - | 0.7 (0.0, 4.7) | - | 0.5 (0.0, 2.8) |
| 10-19 | - | 1.9 (0.2, 6.8) | - | 1.9 (0.2, 6.8) | - | 1.9 (0.2, 6.8) |
| 20-49 | - | ref | - | ref | - | ref |
| 50-64 | - | 1.2 (0.4, 3.6) | - | 1.2 (0.4, 3.6) | - | 1.2 (0.4, 3.6) |
| 65+ | - | 1.9 (0.3, 8.7) | - | 1.9 (0.3, 8.7) | - | 1.9 (0.3, 8.7) |
| **ΔWAIC** | -24.7 |  | -15.1 |  | -15.5 |  |

**Supplemental Table 3.** Attributable fraction of extra-household infections, within household infections by symptomatics, and within household infections by asymptomatics. CrI stands for credible interval.

|  | Model 0;<br>posterior<br>median (95%<br>CrI) | Model 1;<br>posterior<br>median (95%<br>CrI) | Model 2;<br>posterior<br>median (95%<br>CrI) | Model 3;<br>posterior<br>median (95%<br>CrI) | Model 4;<br>posterior<br>median (95%<br>CrI) |
| --- | --- | --- | --- | --- | --- |
| % of all infections from transmission between households members | 22.5 (20.1, 24.2) | 22.5 (20.1, 24.2) | 22.5 (20.1, 24.2) | 22.5 (20.1, 24.2) | 22.5 (20.1, 24.2) |
| % of all infections from transmission between household members by household size |  |  |  |  |  |
| 2 | 23.2 (19.6, 25.9) | 23.2 (19.6, 25.9) | 23.2 (19.6, 25.9) | 23.2 (18.8, 25.9) | 23.2 (19.6, 25.9) |
| 3 | 21.6 (15.7, 23.5) | 19.6 (13.7, 23.5) | 19.6 (15.7, 23.5) | 19.6 (15.7, 23.5) | 19.6 (13.7, 23.5) |
| 4 | 31.2 (26.0, 33.8) | 29.9 (26.0, 33.8) | 29.9 (25.6, 33.8) | 29.9 (26.0, 33.8) | 31.2 (26.0, 33.8) |
| 5 | 41.2 (29.4, 47.1) | 41.2 (29.4, 47.1) | 41.2 (29.4, 47.1) | 41.2 (29.4, 47.1) | 41.2 (29.4, 47.1) |
| % of household infections from asymptomatic individuals | - | - | - | - | 14.5 (7.2, 22.7) |

|  | ***Model 5\*\*;***<br>***posterior***<br>***median (95%***<br>***Crl)*** | Model 6;<br>posterior<br>median (95%<br>Crl) | Model 7;<br>posterior<br>median (95%<br>Crl) | Model 8;<br>posterior<br>median (95%<br>Crl) | Model 9;<br>posterior<br>median (95%<br>Crl) |
| --- | --- | --- | --- | --- | --- |
| % of all household infections from transmission between household members | 22.5 (20.5, 24.5) | 22.5 (20.5, 24.2) | 22.1 (20.1, 24.2) | 22.5 (20.5, 24.2) | 22.5 (20.0, 24.2) |
| % of all household infections from transmission between household members by household size |  |  |  |  |  |
| 2 | 23.2 (19.6, 26.8) | 23.2 (19.6, 25.9) | 23.2 (19.6, 25.9) | 23.2 (19.6, 25.9) | 23.2 (19.6, 25.9) |
| 3 | 19.6 (15.7, 23.5) | 19.6 (13.7, 23.5) | 19.6 (13.7, 23.5) | 19.6 (13.7, 23.5) | 19.6 (13.7, 23.5) |
| 4 | 31.2 (26.0, 33.8) | 31.2 (26.0, 33.8) | 29.9 (26.0, 33.8) | 31.2 (26.9, 33.8) | 29.9 (25.6, 33.8) |
| 5 | 41.2 (29.4, 47.1) | 41.2 (29.4, 47.1) | 41.2 (29.4, 47.1) | 41.2 (29.4, 47.1) | 41.2 (29.4, 47.1) |
| % of household infections from asymptomatic individuals | 14.7 (6.3, 23.2) | 14.5 (7.7, 22.2) | 14.3 (6.4, 22.7) | 13.0 (6.2, 20.3) | - |

\*\* Main model of the study

**Supplemental Table 4.** Estimated probability of infection (%) from extra-household exposures from the start of the epidemic in Geneva until the time of the serosurvey and a single infected household member by age group and sex of the exposed individuals. Graphic representation of the results is shown in Figure 2. CrI stands for credible interval.

|  | Cumulative probability of infection from extra-household sources (95%CrI) |  |  | Household transmission probability from a single infected household member (95%CrI) |  |  |
| --- | --- | --- | --- | --- | --- | --- |
| Age group | All | Male | Female | All | Male | Female |
| 5-9 | 3.6 (1.4, 7.4) | 4.1 (1.6, 8.5) | 3.1 (1.2, 6.2) | 7.6 (1.4, 20.7) | 9.4 (1.7, 26.6) | 6.1 (1.3, 18.1) |
| 10-19 | 3.7 (2.0, 6.1) | 4.4 (2.3, 7.4) | 3.2 (1.7, 5.7) | 15.8 (7.4, 26.1) | 18.0 (8.2, 30.9) | 12.4 (4.7, 23.6) |
| 20-49 | 7.4 (6.0, 9.1) | 8.5 (6.6, 10.7) | 6.4 (4.8, 8.2) | 16.5 (10.3, 24.2) | 20.1 (11.0, 31.6) | 13.9 (6.8, 23.2) |
| 50-64 | 5.2 (4.2, 6.6) | 6.1 (4.7, 8.0) | 4.6 (3.3, 6.1) | 17.4 (9.7, 26.0) | 20.7 (10.5, 33.7) | 14.3 (6.7, 24.6) |
| 65+ | 3.1 (2.2, 4.2) | 3.5 (2.4, 5.0) | 2.6 (1.8, 3.7) | 29.9 (14.0, 49.2) | 36.2 (14.1, 59.6) | 26.1 (11.3, 45.8) |
| All | 5.1 (4.5, 5.8) | - | - | 17.3 (13.7, 21.7) | - | - |

**Supplemental Table 5.** Characteristics of those excluded due to being in an incomplete household to those included in the main analysis.

|  | Study participants in complete households, % (n); N=4,534 | Study participants in incomplete household, % (n); N=3,810 | p-value |
| --- | --- | --- | --- |
| Age |  |  |  |
| 5-9 | 3.7% (167) | 2.8% (106) | <0.0001 |
| 10-19 | 10.1% (459) | 9.0% (344) |  |
| 20-49 | 28.7% (1302) | 42.8% (1632) |  |
| 50-64 | 31.8% (1443) | 32.8% (1251) |  |
| 65+ | 25.7% (1163) | 12.5% (477) |  |
| **Serostatus** |  |  |  |
| seropositive | 6.6% (298) | 7.7% (292) | 0.058 |
| seronegative | 93.4% (4236) | 92.3% (3517) |  |
| **Age specific seropositivity rate** |  |  |  |
| 5-9 | 4.8% (8/167) | 0.94% (1/106) | 0.019 |
| 10-19 | 6.8% (31/459) | 9.9% (34/344) |  |
| 20-49 | 9.1% (119/1302) | 8.8% (144/1632) |  |
| 50-64 | 6.7% (96/1443) | 6.7% (84/1251) |  |
| 65+ | 3.8% (44/1163) | 6.1% (29/477) |  |
| Sex |  |  |  |
| Female | 53.6% (2432) | 53.4% (2033) | 0.82 |
| Male | 46.4% (2102) | 46.6% (1777) |  |
| Self-reported symptoms among the seropositives |  |  |  |
| Symptomatic | 71.5% (213/298) | 75% (219/292) | <0.0001 |
| Asymptomatic | 28.5% (85/298) | 25% (73/292) |  |
| Education Level |  |  |  |
| Compulsory School | 8.5% (385) | 9.4% (357) | 0.051 |
| High School | 16.6% (751) | 17.2% (656) |  |
| Vocational School | 14.0% (637) | 12.6% (481) |  |
| College | 42.2% (1913) | 45.4% (1730) |  |
| Doctorate | 5.1% (229) | 4.6% (176) |  |
| Other | 4.6% (207) | 4.1% (155) |  |
| Missing Response | 9.1% (412) | 6.7% (255) |  |
| Reduced Contact |  |  |  |
| No | 4.9% (224) | 6.0% (229) | 0.055 |
| Yes | 86.3% (3914) | 86.8% (3308) |  |
| Missing Response | 8.7% (396) | 7.2% (273) |  |

**Supplemental Table 6.**
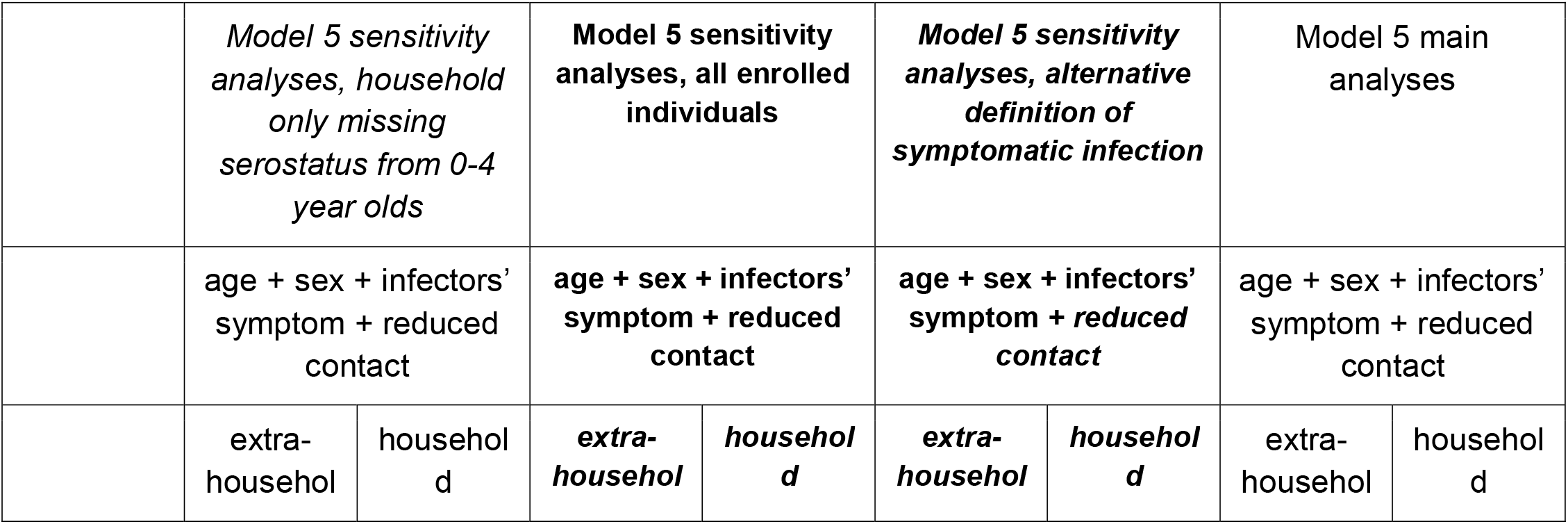
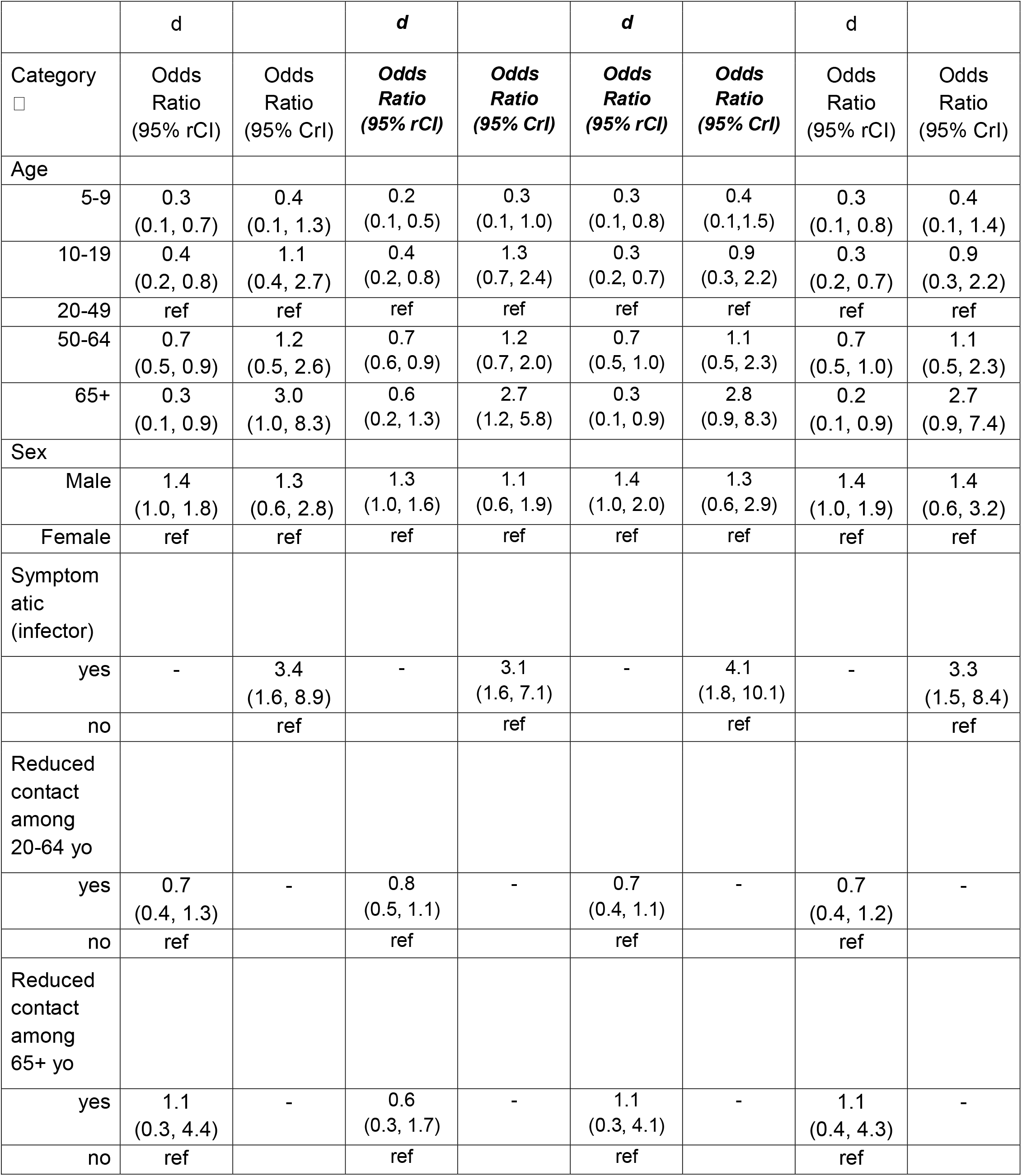
Sensitivity analyses using expanded population and a different definition of symptomatic infection. In the first sensitivity analyses (results shown in the 2nd and 3rd columns), in addition to the 2,267 households included in the main analyses, the study population also included 141 households that are only missing blood samples from household members who are 0-4 years old. In the second sensitivity analyses (results shown in the 4th and 5th columns), the study population included all 8,344 enrolled individuals. In the third sensitivity analyses (results shown in the 6th and 7th columns), we considered seropositive individuals who reported symptom onset within the 2 weeks prior to testing asymptomatic. We ran the main model (model 5) for all three sensitivity analyses. Results of the main model presented in supplemental table 2 are also shown here in the 8th and 9th columns for comparison. The main model incorporates individual-level factors including age, sex, reduced extra-household exposure of the exposed individuals and symptom status of the potential infectors.

### Supplemental Text: Technical summary

#### 1. Chain binomial model description and main assumptions

We built a series of models to estimate two quantities: 1) infection risk from extra-household sources and 2) infection risk from a single infected household member. These models are based on an adapted version of chain binomial models ^14,26^ that we fit to the final size of infections within households.

The model assumes that 1) each household member can be infected either from within a household or from extra-household sources, 2) household members mix at random within a household and can infect one another, and 3) all household members were initially susceptible to infection to SARS-CoV-2, and that infection to SAR-CoV-2 confers immunity to reinfection for the duration of the study period. The explicit modeling of chains of transmission accounts for competing risks between community and household infection. In addition, we assume our serological survey fully captures all infections in a household with no misclassification.

We consider all possible sequences of viral introductions to each household and subsequent transmission events within the household. For example, in a household with 2 seropositive individuals, both could have been infected outside of the household, or one could have been infected outside and then infected one other person within the household. For each possible sequence of viral introduction and subsequent transmission events within the household, we assign generation of infections for each household member (i.e., generation for household member *i*). So, people infected from outside the household are assigned to generation 0, those they infect to generation 1, those generation 1 infects to generation 2, and so on. Uninfected individuals are assigned an implicit generation of infinity. We augment the data and denote each assignment for all members for household *h* as *HH*_*h,k*_, where *k* denotes one possible sequence of viral introduction and subsequent transmission events within the household.

We define the probability of a household member *i* escaping infection from a single infectious household member *j* to be and the probability of individual *i* escaping infection from the community (i.e., outside household contacts) over the course of epidemic to be. We define the probability of household member *i* having an infection generation of as:

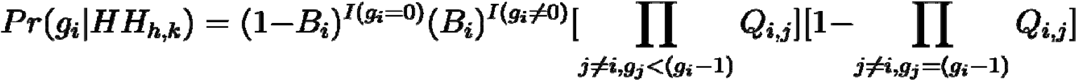

where the first three terms,

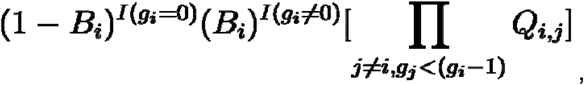

represent the probability of household member *i* escaping infection from extra-household sources and other infected household members up to generation, and

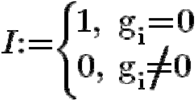

is an indicator function that equals to one if household member i is infected outside.

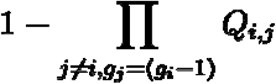

denotes the probability of household member *i* being infected by any infected household member in generation.

We estimate as a function of an exposed individual’s characteristics (i.e., age and sex) and the potential infectors’ characteristics (i.e, symptoms and/or age).

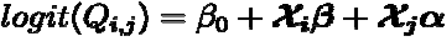

Similarly, we estimate, the probability of household member i escaping infection from extra-household sources since the start of the epidemic, as a function of an exposed individual’s characteristics (age and sex) in addition to two variables capturing self-reported behviours related to social contacts: the reduction in social interactions since the start of the pandemic, and the frequency of extra-household social contacts during the pandemic.

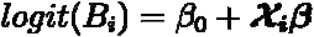

#### 2. Inference

Posterior distributions of parameters are estimated via MCMC using the rstan package, where the likelihood of each observed household pattern of infections is calculated as follows.

The likelihood of each generation assignment is:

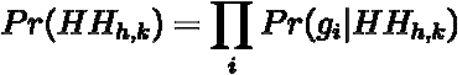

And the likelihood of observing the final infection state (i.e., household attack rate) of a household *h* is then:

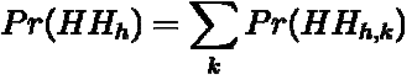

We set weakly informative priors on all parameters to be normally distributed on the logit scale with mean of 0 and standard error of 1.5. We ran four chains of 1,000 iterations each with 250 warm-up iterations and assessed convergence visually and using the Gelman-Rubin Convergence Statistic (R-hat).

#### 3. Simulation of Infectors

We simulate the source of infection for all individuals in the study. We first draw one sequence of viral introductions and subsequent within-household transmission events(*HH*_*h,k*_) from all possible sequences for each household, h, with at least one seropositive individual. Sequences are drawn for each household with probability vector *Pr*(*HH*_*h*,_.) following a categorical distribution,

Next, for each individual, *i*, infected by a household member, we draw the person’s infector from all household members infected in the previous generation. Infectors are drawn from categorical distribution with the probability of each potential infector, j, being 1−*Q*_*i,j*_.

#### 4. Handling of missing variables

As the questions related to extra-household exposures were only asked to those 14 and older, we imputed their responses based on others in their household for analyses that included these variables. To impute the number of extra-household contacts, we took the midpoint of each category (e.g., 4 if answering 3 to 5 contacts per week) and calculated the average response of household members of those missing this variable. We imputed missing behavior pre and post-pandemic with the more common response of household members of those missing this variable. Imputing missing data with the average response of those in the same age and sex category as those missing these two variables did not qualitatively change our estimates.

**Supplemental Table 7.**
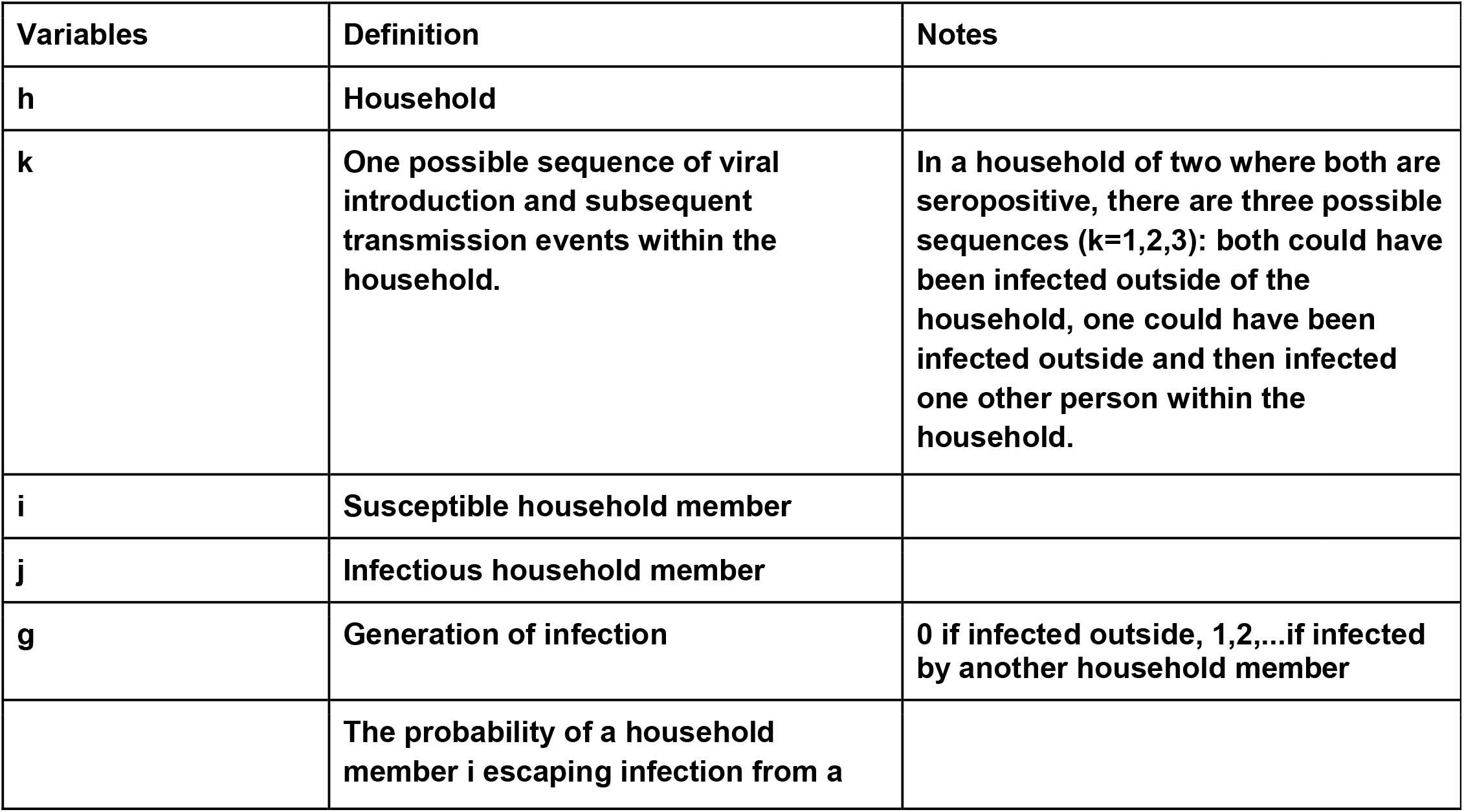
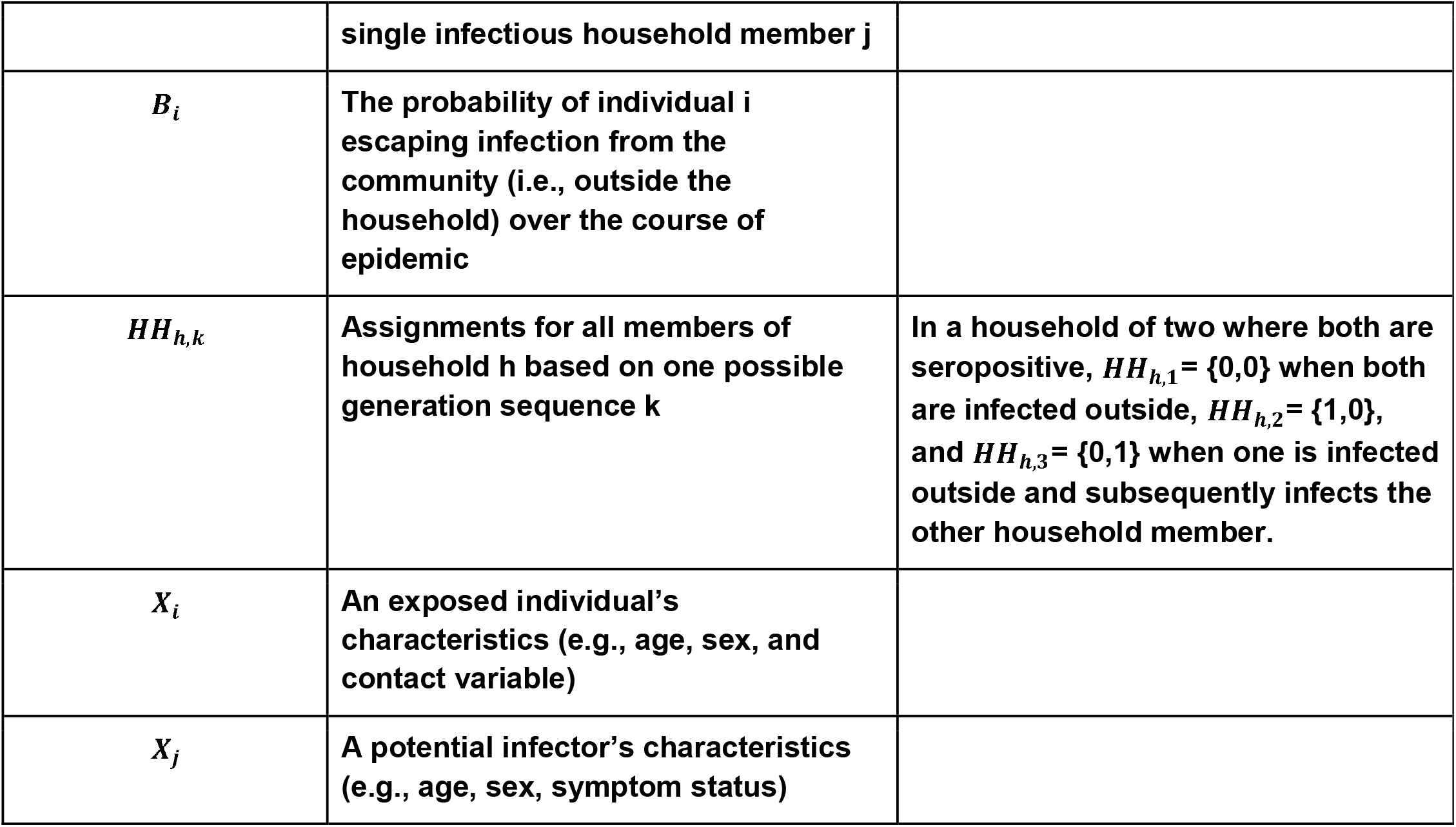
Definition of variables and notations.

| Variables | Definition | Notes |
| --- | --- | --- |
| **h** | **Household** |  |
| **k** | **One possible sequence of viral introduction and subsequent transmission events within the household.** | **In a household of two where both are seropositive, there are three possible sequences (k=1,2,3): both could have been infected outside of the household, one could have been infected outside and then infected one other person within the household.** |
| **i** | **Susceptible household member** |  |
| **j** | **Infectious household member** |  |
| **g** | **Generation of infection** | **0 if infected outside, 1,2,...if infected by another household member** |
|  | **The probability of a household member i escaping infection from a** |  |
|  | single infectious household member j |  |
| $B_i$ | The probability of individual i escaping infection from the community (i.e., outside the household) over the course of epidemic | |
| $HH_{h,k}$ | Assignments for all members of household h based on one possible generation sequence k | In a household of two where both are seropositive, $HH_{h,1} = \{0,0\}$ when both are infected outside, $HH_{h,2} = \{1,0\}$, and $HH_{h,3} = \{0,1\}$ when one is infected outside and subsequently infects the other household member. |
| $X_i$ | An exposed individual's characteristics (e.g., age, sex, and contact variable) | |
| $X_j$ | A potential infector's characteristics (e.g., age, sex, symptom status) | |

**Supplemental Table 8.**
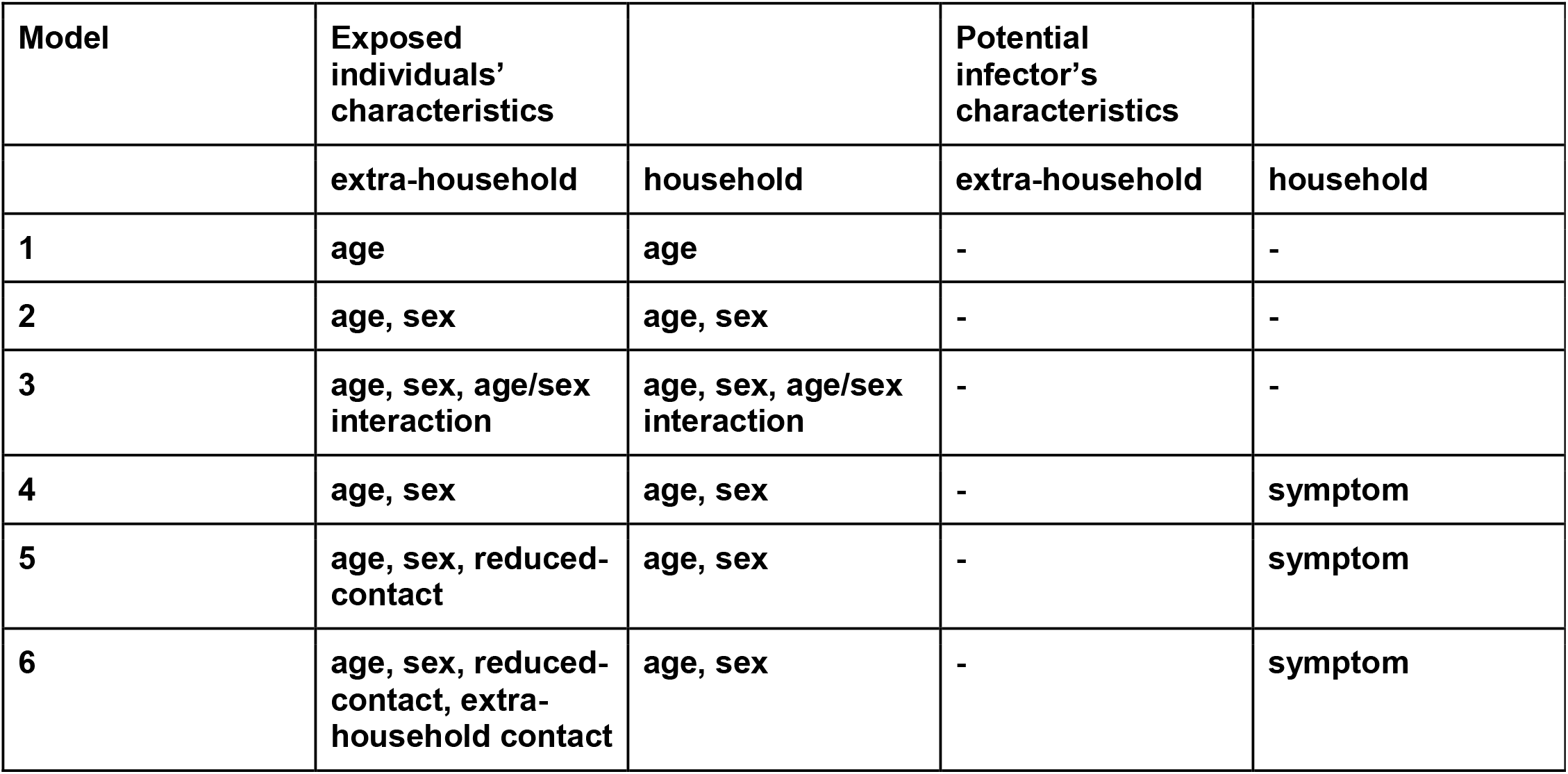
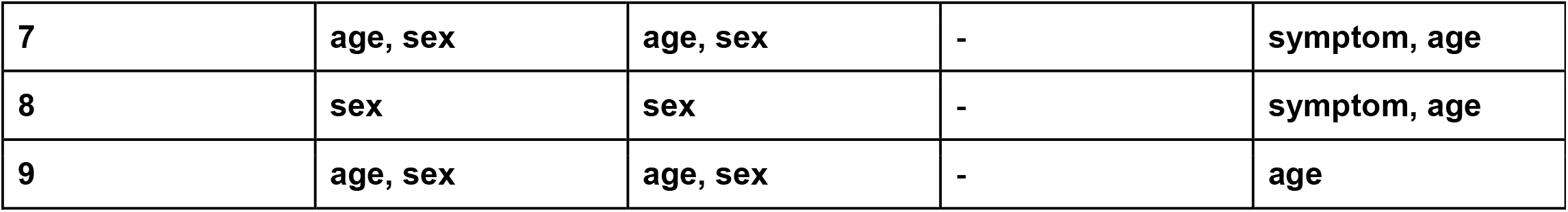
Types of individual characteristics that each model adjusted for for estimating the within household and extra-household transmission risk.

## SEROCoV-POP STUDY TEAM

Silvia Stringhini^1, 2, 3^, Idris Guessous^1, 2^, Andrew S. Azman^1,4,5^, Hélène Baysson^2^, Prune Collombet^1,2^, David De Ridder^2^, Paola d’Ippolito^1^, Matilde D’asaro-Aglieri Rinella^1^, Yaron Dibner^1^, Nacira El Merjani^1^, Natalie Francioli^1^, Marion Frangville^2^, Kailing Marcus^1^, Chantal Martinez^1^, Natacha Noel^1^, Francesco Pennacchio^1^, Javier Perez-Saez^4,5^, Dusan Petrovic^1,3^, Attilio Picazio^1^, Alborz Pishkenari^1^, Giovanni Piumatti^1,8^, Jane Portier^1^, Caroline Pugin^1^, Barinjaka Rakotomiaramanana^1^, Aude Richard^1,4^, Lilas Salzmann-Bellard^1^, Stephanie Schrempft^1^, Maria-Eugenia Zaballa^1^, Zoé Waldmann^2^, Ania Wisniak^4^, Alioucha Davidovic^2^, Joséphine Duc^2^, Julie Guérin^2^, Fanny Lombard^2^, Manon Will^2^, Antoine Flahault^1,2,4^, Isabelle Arm Vernez^9^, Olivia Keiser^4^, Loan Mattera^17^, Magdalena Schellongova^2^, Laurent Kaiser^2,6,9,14^, Isabella Eckerle ^2,6,9^, Pierre Lescuyer^6^, Benjamin Meyer^2, 13^, Géraldine Poulain^6^, Nicolas Vuilleumier^2,6^, Sabine Yerly^6,9^, François Chappuis^1,2^, Sylvie Welker^1^, Delphine Courvoisier^1^, Laurent Gétaz^1,2^, Mayssam Nehme^1^, Febronio Pardo^22^, Guillemette Violot^23^, Samia Hurst^7^, Philippe Matute^1^, Jean-Michel Maugey^22^, Didier Pittet ^12^, Arnaud G. L’Huillier^2,10^, Klara M. Posfay-Barbe^2,10^, Jean-François Pradeau^22^, Michel Tacchino^22^, Didier Trono^11^

1. Division of Primary Care, Geneva University Hospitals, Geneva, Switzerland
2. Faculty of Medicine, University of Geneva, Geneva, Switzerland
3. University Centre for General Medicine and Public Health, University of Lausanne, Lausanne, Switzerland
4. Institute of Global Health, Faculty of Medicine, University of Geneva, Geneva, Switzerland
5. Department of Epidemiology, Johns Hopkins Bloomberg School of Public Health, Baltimore, USA
6. Division of Laboratory Medicine, Geneva University Hospitals, Geneva, Switzerland
7. Institut Ethique, Histoire, Humanités, University of Geneva, Geneva, Switzerland
8. Faculty of BioMedicine, Università della Svizzera italiana, Lugano, Switzerland
9. Geneva Center for Emerging Viral Diseases and Laboratory of Virology, Geneva University Hospitals, Geneva, Switzerland
10. Division of General Pediatrics, Geneva University Hospitals, Geneva, Switzerland
11. School of Life Sciences, Ecole Polytechnique Fédérale de Lausanne (EPFL), Lausanne, Switzerland
12. Infection Prevention and Control program and World Health Organization (WHO) Collaborating Centre on Patient Safety, Geneva University Hospitals, Geneva, Switzerland
13. Centre for Vaccinology, Department of Pathology and Immunology, University of Geneva, Geneva, Switzerland
14. Division of Infectious Diseases, Geneva University Hospitals, Geneva, Switzerland
15. Division of Diagnostics, Geneva University Hospitals, Geneva, Switzerland
16. Division of Women, Children and Adolescents, Geneva University Hospitals, Geneva, Switzerland
17. Campus Biotech, Geneva, Switzerland
18. Education Structure, University of Geneva, Geneva, Switzerland
19. Institute of Social and Preventive Medicine, Bern, Switzerland
20. Deutsches Primatenzentrum (DPZ), Göttingen University, Göttingen, Germany
21. Human Resources Department, Geneva University Hospitals, Geneva, Switzerland
22. Information Systems Division, Geneva University Hospitals, Geneva, Switzerland
23. Division of Communication, Geneva University Hospitals, Geneva, Switzerland

## Notes

### Competing Interest Statement

The authors have declared no competing interest.

### Funding Statement

This study grants from Swiss Federal Office of Public Health, Swiss School of Public Health (Corona Immunitas research program), Fondation de Bienfaisance du Groupe Pictet, Fondation Ancrage, Fondation Privee des Hopitaux Universitaires de Geneve, and Center for Emerging Viral Diseases.

### Author Declarations

The study was approved by the Cantonal Research Ethics Commission of Geneva, Switzerland (CER16-363).

## References

1. Fung HF, Martinez L, Alarid-Escudero F, et al. The household secondary attack rate of SARS-CoV-2: A rapid review. Clin Infect Dis. Published online October 12, 2020. doi:10.1093/cid/ciaa1558

2. Madewell ZJ, Yang Y, Longini IM Jr, Halloran ME, Dean NE. Household Transmission of SARS-CoV-2: A Systematic Review and Meta-analysis. JAMA Netw Open. 2020;3(12):e2031756.

3. Kucirka LM, Lauer SA, Laeyendecker O, Boon D, Lessler J. Variation in False-Negative Rate of Reverse Transcriptase Polymerase Chain Reaction-Based SARS-CoV-2 Tests by Time Since Exposure. Ann Intern Med. Published online May 13, 2020. doi:10.7326/M20-1495

4. Zhang Z, Bi Q, Fang S, et al. Insights into the practical effectiveness of RT-PCR testing for SARS-CoV-2 from serologic data, a cohort study. doi:10.1101/2020.09.01.20182469

5. Iyer AS, Jones FK, Nodoushani A, et al. Persistence and decay of human antibody responses to the receptor binding domain of SARS-CoV-2 spike protein in COVID-19 patients. Sci Immunol. 2020;5(52). doi:10.1126/sciimmunol.abe0367

6. Rosado J, Pelleau S, Cockram C, et al. Multiplex assays for the identification of serological signatures of SARS-CoV-2 infection: an antibody-based diagnostic and machine learning study. The Lancet Microbe. Published online December 21, 2020. doi:10.1016/S2666-5247(20)30197-X

7. Ainsworth M, Andersson M, Auckland K, et al. Performance characteristics of five immunoassays for SARS-CoV-2: a head-to-head benchmark comparison. Lancet Infect Dis. 2020;20(12):1390–1400.

8. Li W, Zhang B, Lu J, et al. The characteristics of household transmission of COVID-19. Clin Infect Dis. Published online April 17, 2020. doi:10.1093/cid/ciaa450

9. Wang Z, Ma W, Zheng X, Wu G, Zhang R. Household transmission of SARS-CoV-2. J Infect. 2020;81(1):179–182.

10. Jing Q-L, Liu M-J, Zhang Z-B, et al. Household secondary attack rate of COVID-19 and associated determinants in Guangzhou, China: a retrospective cohort study. Lancet Infect Dis. Published online June 17, 2020. doi:10.1016/S1473-3099(20)30471-0

11. Richard A, Wisniak A, Perez-Saez J, et al. Seroprevalence of anti-SARS-CoV-2 IgG antibodies, risk factors for infection and associated symptoms in Geneva, Switzerland: a population-based study. bioRxiv. Published online December 18, 2020. doi:10.1101/2020.12.16.20248180

12. Stringhini S, Wisniak A, Piumatti G, et al. Seroprevalence of anti-SARS-CoV-2 IgG antibodies in Geneva, Switzerland (SEROCoV-POP): a population-based study. Lancet. 2020;396(10247):313–319.

13. Meyer B, Torriani G, Yerly S, et al. Validation of a commercially available SARS-CoV-2 serological immunoassay. Clin Microbiol Infect. Published online June 27, 2020. doi:10.1016/j.cmi.2020.06.024

14. Longini IM Jr, Koopman JS. Household and community transmission parameters from final distributions of infections in households. Biometrics. 1982;38(1):115–126.

15. Watanabe S, Opper M. Asymptotic equivalence of Bayes cross validation and widely applicable information criterion in singular learning theory. J Mach Learn Res. 2010;11(12). http://www.jmlr.org/papers/volume11/watanabe10a/watanabe10a.pdf

16. Carpenter B, Gelman A, Hoffman MD, et al. Stan: A probabilistic programming language. J Stat Softw. 2017;76(1). https://www.osti.gov/biblio/1430202

17. Gelman A, Rubin DB. Inference from Iterative Simulation Using Multiple Sequences. Statistical Science. 1992;7(4):457–472. doi:10.1214/ss/1177011136

18. Office fédéral de la statistique. Ménages. Accessed September 17, 2020. https://www.bfs.admin.ch/bfs/fr/home/statistiken/bevoelkerung/stand-entwicklung/haushalte.html

19. Davies NG, Klepac P, Liu Y, et al. Age-dependent effects in the transmission and control of COVID-19 epidemics. Nat Med. 2020;26(8):1205–1211.

20. Kissler SM, Fauver JR, Mack C, et al. Viral dynamics of SARS-CoV-2 infection and the predictive value of repeat testing. Epidemiology. Published online October 23, 2020. doi:10.1101/2020.10.21.20217042

21. Viner RM, Mytton OT, Bonell C, et al. Susceptibility to SARS-CoV-2 Infection Among Children and Adolescents Compared With Adults: A Systematic Review and Meta-analysis. JAMA Pediatr. Published online September 25, 2020. doi:10.1001/jamapediatrics.2020.4573

22. Flasche S, John Edmunds W. The role of schools and school-aged children in SARS-CoV-2 transmission. The Lancet Infectious Diseases. Published online 2020. doi:10.1016/s1473-3099(20)30927-0

23. Salje H, Tran Kiem C, Lefrancq N, et al. Estimating the burden of SARS-CoV-2 in France. Science. 2020;369(6500):208–211.

24. Jääskeläinen AJ, Kekäläinen E, Kallio-Kokko H, et al. Evaluation of commercial and automated SARS-CoV-2 IgG and IgA ELISAs using coronavirus disease (COVID-19) patient samples. Euro Surveill. 2020;25(18). doi:10.2807/1560-7917.ES.2020.25.18.2000603

25. Weidner L, Gänsdorfer S, Unterweger S, et al. Quantification of SARS-CoV-2 antibodies with eight commercially available immunoassays. J Clin Virol. 2020;129:104540.

26. Fraser C, Cummings DAT, Klinkenberg D, Burke DS, Ferguson NM. Influenza Transmission in Households During the 1918 Pandemic. Am J Epidemiol. 2011;174(5):505–514.

